# Menopause in the All of Us Research Program: A Descriptive Summary of Electronic Health Record and Survey Response across Sociodemographic Characteristics

**DOI:** 10.64898/2026.04.17.26351129

**Authors:** Jack W. Staples, Samantha L. White, Adam Giacalone, Nikita Pozdeyev, Mary D. Sammel, Barbara E. Stranger, Celina I. Valencia, Nanette Santoro, Audrey E. Hendricks

## Abstract

**Objective:** Menopause is a significant physiological transition with implications for health outcomes (e.g., cardiometabolic), yet gaps remain in understanding the menopause transition, including how menopause timing and type influence health outcomes. Large-scale cohort studies in midlife (age∼40-60) females, including the All of Us Research Program (AoURP), provide opportunities to study menopause across diverse populations and data modalities. We characterized menopause-related data in AoURP, focusing on age distributions and concordance between EHR diagnosis codes and self-reported survey responses.

**Methods:** We analyzed menopause-related survey, EHR diagnostic code, and genomic data among ∼396,000 participants in AoURP with female sex. We summarized menopause data across modalities, overlap between survey, EHR, and genomic data, and age distributions overall and across sociodemographic characteristics.

**Results:** Among ∼396,000 females, surveys captured ∼193,000 menopause observations, nearly seven times more than structured EHR diagnoses (∼28,000), suggesting under-ascertainement in EHR data. Nearly all females (∼99%) with an EHR menopause diagnosis also reported menopause in the survey. Approximately 22,000 participants had intersected EHR, survey, and genomic menopause-related data. Survey-based age patterns matched expectations, with participants <40 years predominantly reporting pre-menopausal status and those >60 years predominantly reporting post-menopausal status. A small subset (N≈1,700; 4%) (age>70 years) reported no menopause, suggesting response or recall bias. EHR menopause codes were concentrated after age>45 years, with a notable spike at age 65. Modest differences in survey-based menopause age distributions were observed by sociodemographic characteristics (e.g., race, ancestry).

**Conclusions:** These findings inform sampling strategies, power calculations, phenotype definition, and study design for menopause research using AoURP.

## INTRODUCTION

Menopause is a significant and variable transition in female physiology accompanied by substantial hormonal shifts [e.g., elevated follicle-stimulating hormone (FSH), decreased estradiol]^1^. While menopause is a single point-in-time occurrence defined as 12 months without a menstrual bleed, the menopause transition is commonly categorized into three stages: pre-, peri-, and post-menopause^1^. Although the age of natural menopause timing varies across individuals, the median age of occurance is ∼51-52 years^2^. Age alone does not determine transition stage or symptom (e.g., vasomotor) presence, but it offers a general framework for categorization^1–3^. Additionally, menopause is associated with increased risk for multiple diseases and conditions, most notably cardiovascular and metabolic (CVM) disease and osteoporosis^4–6^.

Despite its clinical significance, menopause status is often not accounted for when modeling clinical outcomes, limiting the ability to assess symptom variation, treatment response, or health risks associated with the menopause transition^7^. Further, menopause is often inconsistently measured, with missing or incomplete data. This is especially common for electronic health records (EHRs), and can also occur in survey responses^7–9^. In addition to smaller, gold standard studies of women in midlife and menopause, such as the Study of Women’s Health Across the Nation (SWAN), large, publicly available cohorts with menopause information and multimodal data, such as the UK Biobank,enable evaluation of the role of menopause in health and disease at scale^10–12^. The All of Us Research Program (AoURP), a National Institutes of Health (NIH) precision medicine initiative, provides a unique opportunity to study menopause in a large, diverse, publicly available, multimodal US cohort^13,14^.

The AoURP was designed to capture participants across a broad range of sociodemographic, geographic, and health characteristics in the US^13,14^. With >470,000 participants with EHR data in v8 (∼608,000 biosamples collected) and the goal of >1 million contributing a variety of multimodal data, including EHR, survey, genomic, wearables, and physical measurements, the AoURP holds substantial potential to examine menopause status and related characteristics across multiple data modalities. A foundational descriptive analysis can support the broad use of AoURP menopause data in health and disease research by guiding study design and informing power and sampling considerations.

Sociodemographic characteristics are important to consider in menopause research due to their relationship with the timing and symptom severity of the menopause transition^3,15^. Differences in the menopause transition experience are likely driven by a combination of biological (e.g., genomic), environmental, social, economic, and healthcare access factors^16^. A previous study showed that females identifying as Black, Hispanic, Middle Eastern, or more than one ethnicity experienced greater menopause symptom severity, whereas Asian and South Asian participants reported lower symptom severity compared to non-Hispanic White participants^15^.

Additionally, the relationship between menopause and adverse health outcomes often differs by sociodemographic characteristics (e.g., race, ethnicity, socioeconomic status, and lifestyle factors)^3,15^. Thus, incorporating sociodemographic factors into menopause research in large cohort studies is crucial for ensuring findings are relevant to individuals from diverse backgrounds and that analyses reflect real world variability in health experiences.

Here, we present a comprehensive descriptive summary of menopause data across data modalities and sociodemographic strata within the AoURP Controlled Tier v8 and v7 (Supplemental Digital Content 1-2) datasets. Specifically, we (1) report summary statistics for EHR and survey-derived menopause descriptors, (2) evaluate overlap of menopause descriptors and other data types, including genomic data, and (3) assess menopause characteristics across sociodemographic factors. Overall, this study can inform study design, hypothesis generation, and modeling strategies for conducting comprehensive and representative menopause research using AoURP data.

## METHODS

### All of Us Research Program

We conducted analyses using the AoURP Controlled Tier dataset (v8 included in the main text and v7 in Supplemental Digital Content 1-2). We performed data extraction, cleaning, descriptive summary statistics calculation, and data visualization using *structured query language* (*SQL*)^17^, *R* statistical computing language (v4.4.0)^18^, and *Python* (v3.10.16;^19^) within the AoURP workbench [workspaces: “(VERSION 8)” and “(VERSION 7) Duplicate of Genetic Substructure in the All of Us Biobank”] with *tidyverse*^20^, *bigrquery*^21^, *scales*^22^, and *grid* (included as part of base R installation^18^) packages. Throughout the text, sample sizes ≤ 20 are obscured and reported as “N ≤20” or “20” in accordance with the AoURP Data Dissemination Policy. Further, sample sizes >20 are rounded up to the nearest value of 5 to prevent derivation of sample sizes ≤20.

### Sociodemographic variables

AoURP sociodemographic data were sourced from demographic data (date of birth, ethnicity, race, sex) and basic survey (education level: *“highest grade or year of school completed”* and income level: *“annual household income from all sources”*) datasets. For some analyses, we collapsed income levels into fewer groups: $35,000 (35k) or less (<10k, >10k to 25k, >25k to 35k), 35k to 100k (>35k to 50k, >50k to 75k, >75k to 100k), 100k or more (>100k to 150k, >150k to 200k, >200k). Data for individuals of female were defined based on the demographic survey sex at birth variable. We calculated <u>age</u> as the difference between the date of observation (EHR or survey) and date of birth, and then grouped participants into age quartiles [Q1 (age 19–41 years), Q2 (42–57), Q3 (58–69), and Q4 (≥70)] and menopause transition age groups (age <40, 40-60, and >60 years). These age groups approximately correspond to pre-, peri-, and post-menopausal stages, respectively. We chose these age groups to capture as many natural menopause observations as possible because there is known variation around the age of natural menopause (median ∼51-52 years)^2^. Age <40 was considered premenopausal because natural menopause prior to age 40 is rare (∼1% of the US population) and is often considered a distinct clinical entity^23^. Perimenopause is a variable stage that can last 10 years or more, and natural menopause after age 60 is extremely rare^2^. Thus our pre- and post-menopause groups are expected to contain >99% of female participants who are truly pre- or post-menopausal, while the peri-menopausal group represents a mixture of transition states^2,24,25^. Access to individuals was not available and clinical confirmation of pre-, peri-, or post-menopause status was not performed.

### EHR menopause descriptors

We defined <u>EHR menopause diagnostic codes</u> using the Systematized Nomenclature of Medicine Clinical Terms (SNOMED CT) vocabulary, indicating clinical diagnosis of menopause and related conditions. We queried menopause-related SNOMED codes based on their earliest date of appearance in the AoURP EHR Conditions dataset including: *abnormal vasomotor function* (SNOMED 70670009), *menopause ovarian failure* (237138004), *menopause present* (SNOMED 289903006), *premature menopause* (SNOMED 373717006), *premature ovarian failure* (237788002), and *primary ovarian failure* (SNOMED 65846009). Age was calculated at the earliest date of code appearance. Additionally, we counted the number of occurances of the *menopause present* (SNOMED 289903006) EHR code per individual.

### Survey menopause descriptors

We obtained <u>survey menopause data</u> from the women’s health questionnaire from the AoURP Overall Health survey (described in detail at the links in these citations:^26,27^). Questions analyzed included:

(1) “*Q: Have your menstrual periods stopped permanently?*” with response options [*“periods have not stopped”, “yes none”, “yes but hormone”, “not sure Have your menstrual periods stopped permanently?”, “prefer not to answer”, or skip question*];
(2) “*Q: If yes to Have your menstrual periods stopped permanently?, why did your periods stop?*” with response options [“*natural menopause”, “surgery”, “endometrial ablation”, “medication therapy”, “other”, “not sure”, “prefer not answer”, or* skip question];
(3) “*Q: Have you ever had a hysterectomy, that is surgery to remove your uterus or womb?*” with response options [“*no”, “yes”, “not sure”, “prefer not answer”, “no matching concept”, or* skip question];
(4) “*Q: Have you ever had an ovary removed?*” with response options [“*no”, “yes sectioned” (partial), “yes both”, “yes unsure”, “not sure”, “prefer not answer”, or* skip question]. Question responses were selected based on their earliest date of appearance, and age was calculated corresponding at that timepoint.

### Data set creation

To create a combined EHR and survey dataset, we first identified participants with available sociodemographic data using the AoURP Cohort Builder. We then created a second cohort of participants with any menopause-related data from EHR condition codes or survey responses. We joined the sociodemographic, survey, and the menopause EHR condition datasets, encoding diagnostic codes as “1” (presence) or “0” (absence) to generate a combined dataset (i.e., the union of data tables from each modality for each individual). We used the combined dataset to calculate descriptive statistics for clinical EHR- and survey-derived menopause variables across sociodemographic strata.

### Intersections of menopause descriptors from EHR, survey, and genomics

To evaluate overlap in menopause information across data sources, we examined combinations of menopause variables from EHR diagnostic codes, survey responses, and menopause transition age groups (<40, 40-60, and >60 years). Definitions of these variable-level descriptors are provided in the “EHR menopause descriptors” and “Survey menopause descriptors” sections. We also assessed broader patterns of overlap by identifying shared participants across EHR, survey, and genomic datasets. Using unique participant identifiers, we calculated sample sizes for marginal and intersecting sets across EHR, survey, and short read whole-genome sequencing (srWGS) data for: (1) all participants with female sex (defined by sex at birth demographic variable), and (2) participants with female sex and menopause-specific data from these sources, including EHR diagnoses [*menopause present* (SNOMED 289903006) or *premature menopause* (SNOMED 373717006)], and survey responses to the “*Have your menstrual periods stopped permanently?*” question.

### Calculation of descriptive summary statistics

To characterize the distribution of menopause variables for EHR diagnostic codes and survey responses, we calculated sample sizes and the median and interquartile range (IQR) for age overall and within sociodemographic strata.

## RESULTS

### Survey responses identify approximately seven times more menopause events than EHR data

The AoURP v8 cohort includes N≈395,990 female participants with available demographic data (**Tables 1-2, S1-S2**). Menopause was reported for approximately seven times more individuals in self-reported survey responses than in EHR diagnostic codes (**Table 3,S3; Figure 1,S1**). A total of N≈192,655 participants answered “yes” to the survey question, whereas only N≈27,975 participants had at least one menopause diagnostic code in the EHR [N≈27,165 with *menopause present* (SNOMED 289903006); N≈1,220 with *premature menopause* (SNOMED 373717006); N≈415 with both EHR codes]. Almost all individuals identified as menopausal in the EHR were also identified in the survey (>99%, **Figure 1, S1**).

**Figure 1.**
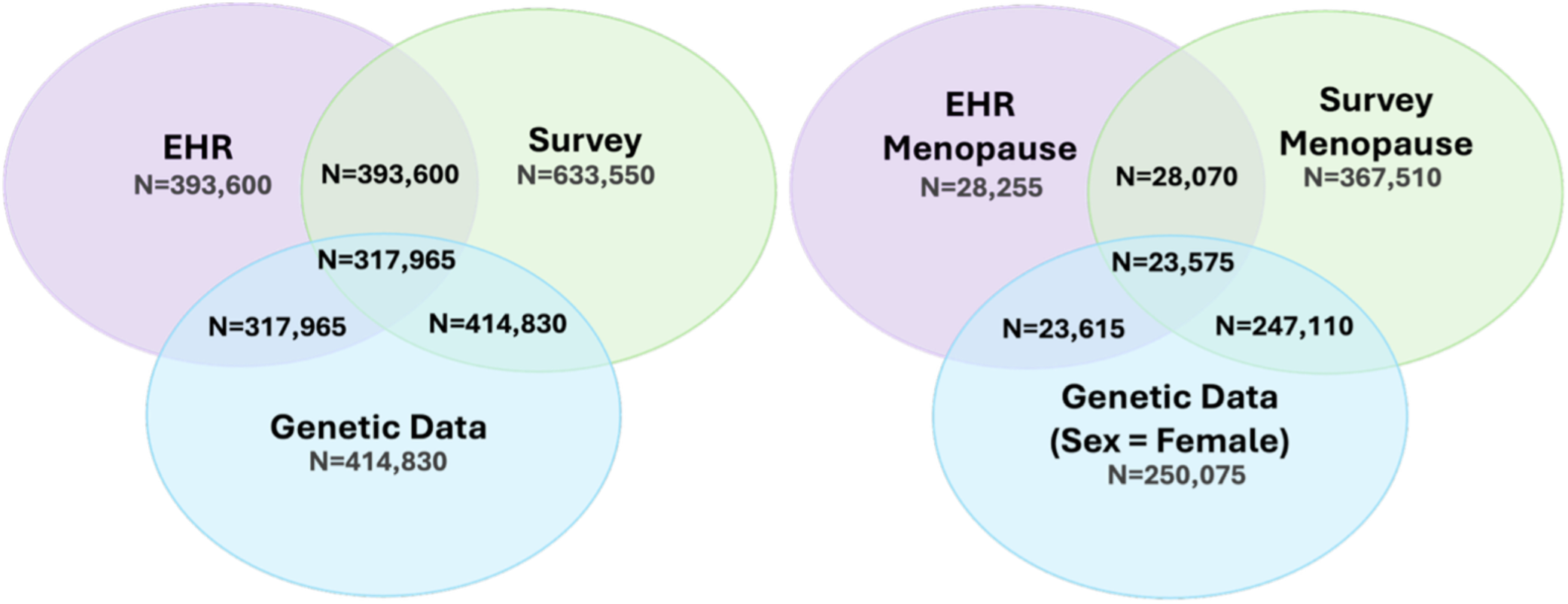
Intersections of EHR, survey, and genomic data among AoURP v8 participants. Venn diagram illustrating intersection sample sizes of participants with EHR, survey, and genomic data for all participants **(left)** and for menopause-related variables among particpants with female sex **(right)**. The EHR Menopause group includes unique participants with at least one SNOMED diagnostic code for *menopause present* (SNOMED 289903006) or *premature menopause* (SNOMED 373717006). The Survey Menopause group includes participants who had any response to the question “*Have your menstrual periods stopped permanently?*”. The Genetic Data (Sex = Female) group includes female participants with short-read whole-genome sequencing (srWGS) data. All participant counts were rounded up to the nearest multiple of 5.

**Table 1.** Sample sizes and age distributions for sociodemographic and menopause variables in the AoURP cohort (participants with sex = female).

| Sociodemographic strata group | N (% of group) | Age (years) summary<sup>a</sup> |  |  |  |
| --- | --- | --- | --- | --- | --- |
|  |  | Q1 | Median | Q3 | IQR |
| **Full AoURP (sex = female)** | 395990 (100%) | 35 | 50 | 63 | 28 |
| **Age quartile** |  |  |  |  |  |
| Q1 (19-41 years) | 141645 (36.0%) | 26 | 31 | 36 | 10 |
| Q2 (42-57 years) | 113140 (28.7%) | 46 | 50 | 54 | 8 |
| Q3 (58-69 years) | 90380 (23.0%) | 60 | 63 | 66 | 6 |
| Q4 (≥70 years) | 48370 (12.3%) | 72 | 74 | 78 | 6 |
| **Age (menopause transition)** |  |  |  |  |  |
| <40 years | 130400 (32.9%) | 25 | 30 | 35 | 10 |
| 40-60 years | 150665 (38.0%) | 45 | 51 | 56 | 11 |
| >60 years | 114925 (29.0%) | 64 | 68 | 73 | 9 |
| **Education level (EL)** |  |  |  |  |  |
| EL1 (never attend) | 480 (0.1%) | 48 | 60 | 69 | 21 |
| EL2 (grade 1-4) | 2980 (0.8%) | 48 | 58 | 67 | 19 |
| EL3 (grade 5-8) | 7615 (1.9%) | 43 | 53 | 63 | 20 |
| EL4 (grade 9-11) | 17990 (4.5%) | 34 | 47 | 57 | 23 |
| EL5 (grade 12, GED) | 64875 (16.4%) | 32 | 47 | 60 | 28 |
| EL6 (college 1-3 years) | 111735 (28.2%) | 35 | 49 | 62 | 27 |
| EL7 (college graduate) | 96515 (24.4%) | 33 | 49 | 62 | 29 |
| EL8 (advanced degree) | 86330 (21.8%) | 38 | 53 | 66 | 28 |
| prefer not to answer, skip | 7495 (1.9%) | 36 | 50 | 61 | 25 |
| **Ethnicity** |  |  |  |  |  |
| Hispanic or Latino | 74905 (18.9%) | 30 | 42 | 55 | 25 |
| not Hispanic or Latino | 311430 (78.6%) | 36 | 51 | 64 | 28 |
| prefer not to answer, skip | 9655 (2.4%) | 39 | 54 | 66 | 27 |
| **Income level (IL)** |  |  |  |  |  |
| IL1 (<10k) | 44220 (11.2%) | 30 | 43 | 56 | 26 |
| IL2 (10k-25k) | 45890 (11.6%) | 35 | 51 | 63 | 28 |
| IL3 (25k-35k) | 31495 (8.0%) | 31 | 46 | 63 | 32 |
| IL4 (35k-50k) | 35950 (9.1%) | 32 | 47 | 64 | 32 |
| IL5 (50k-75k) | 47140 (11.9%) | 35 | 50 | 65 | 30 |
| IL6 (75k-100k) | 35640 (9.0%) | 37 | 52 | 65 | 28 |
| IL7 (100k-150k) | 41910 (10.6%) | 38 | 51 | 63 | 25 |
| IL8 (150k-200k) | 19660 (5.0%) | 38 | 50 | 61 | 23 |
| IL9 (>200k) | 23500 (5.9%) | 40 | 51 | 61 | 21 |
| prefer not to answer, skip | 70605 (17.8%) | 35 | 52 | 64 | 29 |
| **Race** |  |  |  |  |  |
| American Indian Alaska Native | 5120 (1.3%) | 32 | 44 | 56 | 24 |
| Asian | 13305 (3.4%) | 27 | 38 | 53 | 26 |
| Black African American | 59250 (15.3%) | 36 | 50 | 60 | 24 |
| Middle East North Africa | 1905 (0.5%) | 29 | 40 | 56 | 27 |
| more than one population | 20305 (5.1%) | 31 | 43 | 57 | 26 |
| Native Hawaii Pacific Island | 395 (0.1%) | 31 | 43 | 55 | 24 |
| White | 224725 (56.7%) | 37 | 53 | 65 | 28 |
| prefer not to answer, skip | 70995 (17.9%) | 32 | 45 | 58 | 26 |
| **Ancestry (requires genetic data)** |  |  |  |  |  |
| AFR-like | 48655 (19.5%) | 36 | 50 | 60 | 24 |
| AMR-like | 51745 (20.7%) | 30 | 43 | 56 | 26 |
| EAS-like | 6325 (2.5%) | 28 | 40 | 57 | 29 |
| EUR-like | 139595 (55.8%) | 39 | 56 | 67 | 28 |
| MID-like | 820 (0.3%) | 28 | 40 | 56 | 28 |
| SAS-like | 2945 (1.2%) | 26 | 36 | 50 | 24 |
| **Menopause** |  |  |  |  |  |
| EHR [*premature or menopause present*]<sup>b</sup> | 27975 [7.1% of all females (N=395990)] | 59 | 65 | 70 | 11 |
| Survey [*yes for any reason*]<sup>c</sup> | 192655 [48.7% of all females (N=395990)] | 54 | 61 | 69 | 15 |

**Abbreviations:** AoURP, All of Us Research Program; EHR, electronic health record; EL, education level; IL, income level; IQR, interquartile range; N, number of participants; Q1, first quartile; Q3, third quartile. For education and income level, “prefer not to answer” or “skip” responses were combined. For ethnicity, “none of these”, “prefer not to answer”, and “skip” responses were combined. For race, “None Indicated”, “None of these”, “prefer not to answer” and “skip” responses were combined. Participant counts ≤20 are reported as “N ≤20” in accordance with the AoURP Data Dissemination Policy. All other participant counts were rounded up to the nearest multiple of 5. <sup>a</sup>Age summary statistics include Q1, median, Q3, and IQR, calculated from date of birth and the demographic basic survey date; age for menopause descriptors was calculated from date of birth and the earliest observation date in the EHR or survey.
<sup>b</sup>EHR menopause diagnostic codes: *premature menopause* (SNOMED 373717006) or *menopause present* (SNOMED 289903006). <sup>c</sup>Survey menopause: participants who answered “yes” for any reason to the question “Have your menstrual periods stopped permanently?”.

**Table 2.**
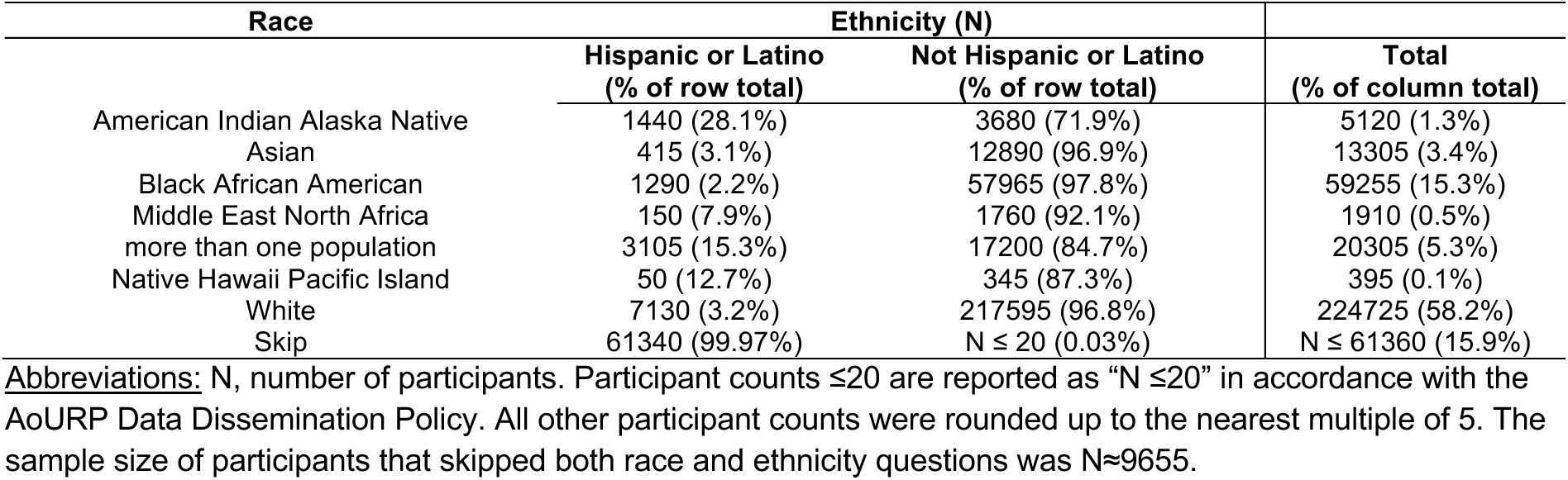
Sample sizes of AoURP participants with female sex across intersections of self-reported race and ethnicity.

**Table 3.**
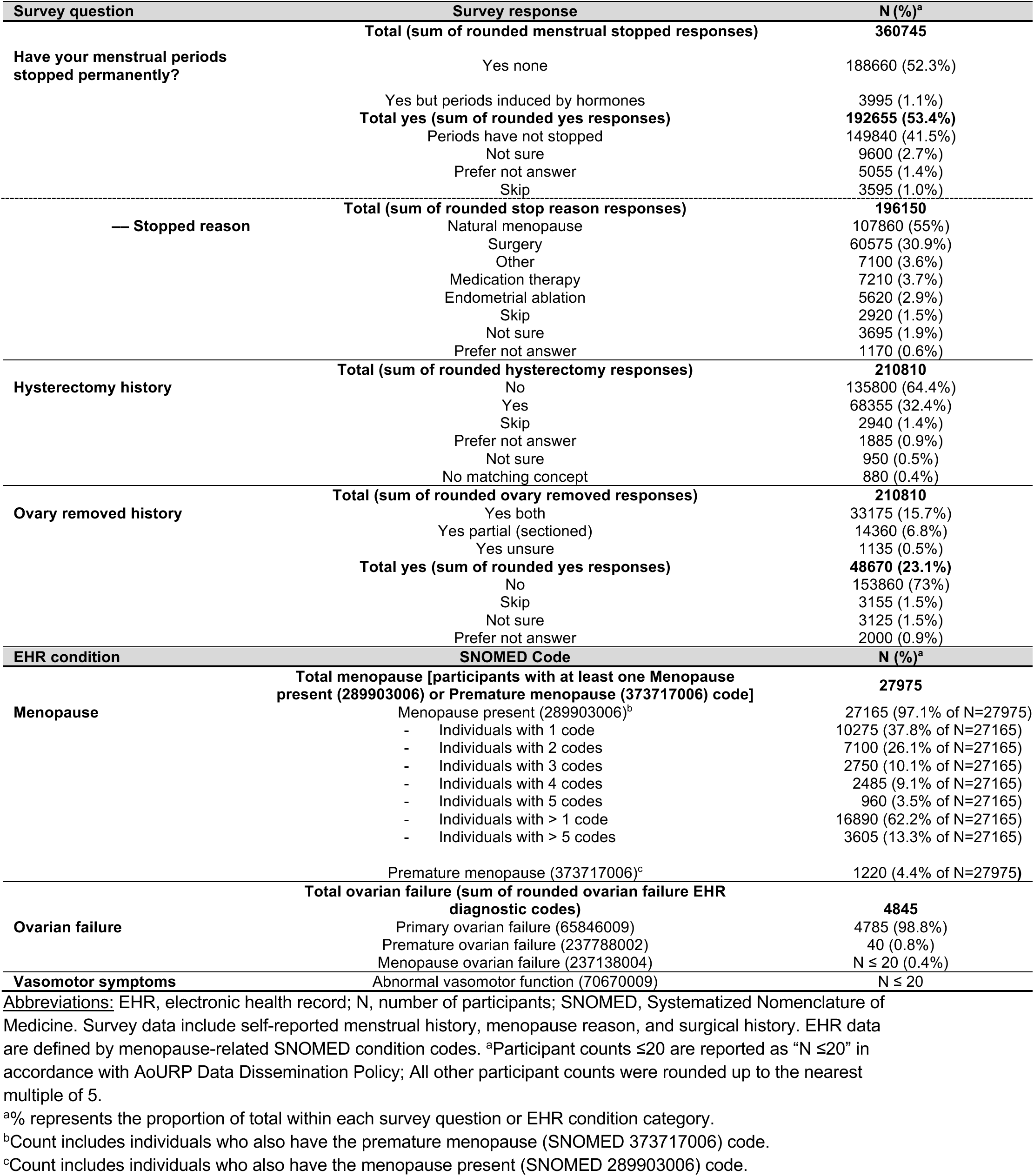
AoURP variable-level survey and EHR-derived diagnostic descriptors of menopause for participants with female sex.

**Tables 3 and S3** highlight nested survey responses and EHR observations of menopause descriptors . For the survey question “*Have your menstrual periods stopped permanently?*”, most participants selected “*yes none*” or “*yes but hormone*” (∼53%, N≈192,655), or “*periods have not stopped*” (∼42%, N≈149,840). Ambiguous responses were less common (combined “*not sure*”, “*prefer not to answer*”, and “*skip*”; N≈18,250; ∼5%). Surgical history responses occurred frequently in the survey (∼31%, N≈60,575). Among individuals reporting “yes” to any “*ovary removed history*” (N=48,670), removal of both ovaries was most common (N≈33,175; ∼68%), with partial (∼30%) or uncertain (∼2%) removal less frequent. In the EHR, menopause diagnoses were often recorded repeatedly for the same individual (N≈16,890 with >1 code versus N≈10,275 with a single code; N≈7,100 with 2 codes; N≈2,750 with 3 codes; N≈2,485 with 4 codes; N≈960 with 5 codes; N≈3,605 with > 5 codes). Within ovarian-failure EHR codes, *primary ovarian failure* (SNOMED 65846009) (N≈4,785) comprised nearly all ovarian failure observations (>99%), indicating focused use of a single SNOMED concept for ovarian failure. Vasomotor symptoms, a common occurrence during the menopause transition, were almost entirely absent from structured EHR data, with N ≤20 observations for *abnormal vasomotor function* (SNOMED 70670009) highlighting a major gap in menopause symptom-level documentation.

### Limited documentation of menopause in the EHR is unlikely to be explained by differences in menopause type

To evaluate consistency between EHR codes and survey responses, we identified the 10 most frequent combinations of menopause descriptors for female participants most likely to be post menopause, age >60. (**Table 4**). Due to the low number of participants with EHR menopause codes, eight of the ten most common combinations did not include menopause identified in the EHR. Of the N≈53,505 participants with the most common survey combination of non-surgical *“natural menopause”* with or without EHR *menopause present* (SNOMED 289903006), only N≈9,565 (∼17.9%) had an EHR code indicating *menopause present* (SNOMED 289903006) (**Table 4**, **Figure 2,S2)**. A similar proportion (18.1%; N≈3,335 of 18,405) had the EHR code indicating *menopause present* (SNOMED 289903006) for the surgical menopause with both a hysterectomy and ovary removal survey response (“*yes both*”, “*yes partial*”, or “*yes unsure*”) (**Table 4**, **Figure 2,S2)**. This suggests that the lack of menopause in the EHR likely reflects systematic under-documentation rather than differences by menopause type or discordant information between data sources. We also note that the survey answer combinations identified in **Table 4** are clinically plausible, such as surgical practices for benign conditions (e.g., hysterectomy for fibroids), hysterectomy with sparing the ovaries, and ovary removal alone for high genetic risk cancer prevention. There are three survey combinations that likely represent surgeries occurring after natural menopause where the survey response of *“natural menopause”* was listed along with *“hysterectomy”* and *“yes”* to both ovary removal (N≈2,980), only hysterectomy (N≈1,470), or only sectioned/partial ovary removal (N≈1,975) (**Table 4**, **Figure 2,S2)**.

**Figure 2.**
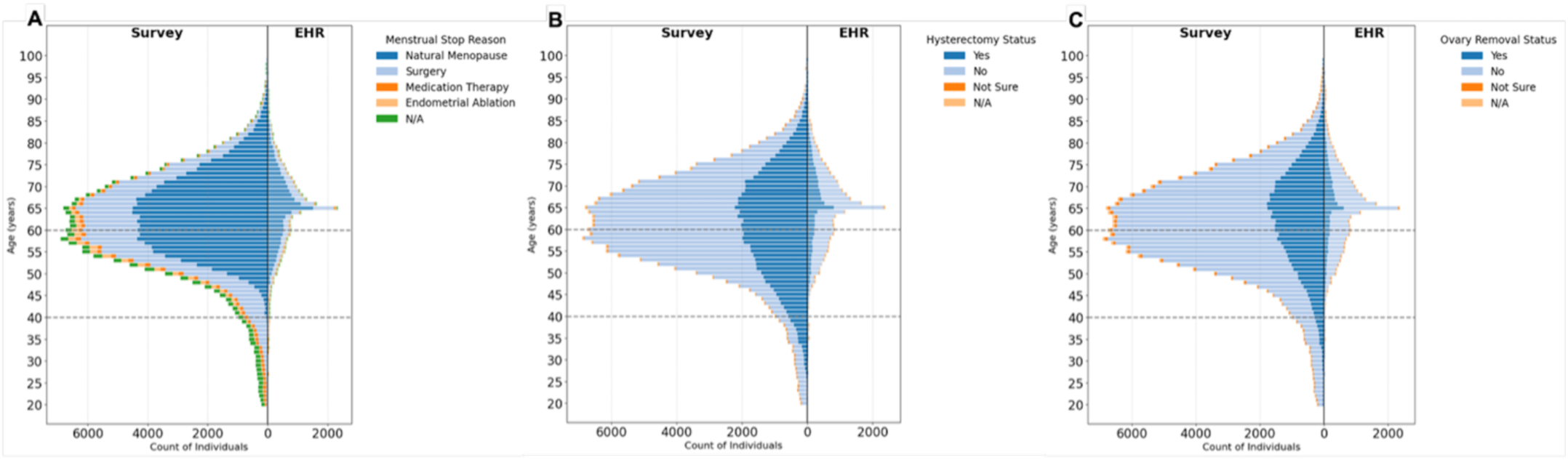
AoURP v8 age distributions in survey and EHR data for menopause classifications. Histograms (1-year bins) are shown for participants with female sex at birth for survey (answered the question “*Have your menstrual periods stopped permanently?*”) **(left panels)** vs. EHR menopause status [SNOMED diagnostic code for *menopause present* (SNOMED 289903006)] **(right panels)** for **(A)** survey reason for menopause (*“menstrual stop reason”*), **(B)** survey “*hysterectomy history*” status, and **(C)** survey “*ovary removal history*” status. Grey dashed lines indicate the approximate menopause transition age range (40-60 years). Sample sizes ≤20 are obscured and reported as “20” in accordance with AoURP Data Dissemination Policy. All other participant counts were rounded up to the nearest multiple of 5.

**Table 4.** Ten most common combinations of menopause reponses across EHR and survey data among participants with female sex aged >60 years.

| N | overall health, women's health survey question and response |  |  |  | EHR SNOMED code |
| --- | --- | --- | --- | --- | --- |
|  | "menstrual stop" | "stop reason" | "hysterectomy" | "an ovary removed" | Menopause present (289903006) |
| 43940 | yes none | natural menopause | no | no | no |
| 11580 | yes none | surgery | no response | yes both | no |
| 9565 | yes none | natural menopause | no | no | yes |
| 6625 | yes none | surgery | yes | no | no |
| 2980 | yes none | natural menopause | yes | yes both | no |
| 2615 | yes none | surgery | yes | yes sectioned/partial | no |
| 2465 | yes none | surgery | yes | yes both | yes |
| 1975 | yes none | natural menopause | no | yes sectioned/partial | no |
| 1905 | periods have not stopped | NA (not stopped) | no response | no response | no |
| 1470 | yes none | natural menopause | yes | no | no |

*menopause present* (SNOMED 289903006) EHR diagnosis; survey responses to the questions “*Have your menstrual periods stopped permanently?*”, “*Have you ever had a hysterectomy, that is surgery to remove your uterus or womb?*”, and “*Have you ever had an ovary removed?*” ; and menopause transition age category. Abbreviations: EHR, electronic health record; SNOMED, Systematized Nomenclature of Medicine; N, number of participants; NA, not applicable. All participant counts were rounded up to the nearest multiple of 5.

### Intersection of EHR, survey, and genomic data highlights potential for integrated analyses

To assess the potential to study menopause using the large genomic AoURP resource, we identified the intersections of participants with menopause variables across EHR, survey, and genomic (srWGS) data (**Figure 1, S1**). Among female participants with genomic data (N≈250,075), ∼9% (N≈23,615) had menopause EHR data, compared to ∼99% with menopause survey responses (N≈247,110). Consequently, ∼9% of female participants with genomic data had both EHR and survey menopause data. Across sociodemographic strata, the sample sizes were lower in the three-way intersection compared to menopause variables from the survey alone due to the limited number of individuals with menopause information in the EHR compared to the survey (**Figure S3A, S4A**).

### Menopause and age patterns in the survey

Among female participants answering “*yes*” to the survey question *“Have your menstrual periods stopped permanently?”* (N=192,655), most attributed menopause to natural causes (N≈107,860; ∼56.0%). Approximately ∼31% cited surgery, while ∼7% reported medication therapy or endometrial ablation (**Table 3,S3; Figure 2,S2**). Female participants who answered “*yes none*” (median=61 years, Q1=54, Q3=69; N≈188,660) to the menopause survey question were typically older than the overall AoURP female cohort (median=50 years, Q1=35, Q3=63; N≈395,990), and females that answered “*yes but hormone*” (median=35 years, Q1=27, Q3=46; N≈3,995) were younger (**Table 1**,**3,S1,S3**). Few “*yes none*” menopause question respondents were aged <40 years (4.6%, N≈8,615), with most aged 40–60 years (42.0%, N≈79,265) or >60 years (53.0%, N≈100,785), consistent with biological expectations. Similarly, “*natural menopause*” without hysterectomy and ovary removal was most frequent in the >60 group (59.9%, N≈55,740) and 40–60 group (39.6%, N≈36,800), and rare in those aged <40 (0.5%, N≈440) (**Table S4-S5 in Supplemental Digital Content 2**).

### Age distributions differ between EHR- and survey-derived menopause data

Unlike the bell-shaped age distribution observed for survey menopause “*yes*” responses, the age distributions for the EHR menopause data were flatter with most data occurrences between ages 50-80. Notably, there is a sharp peak at age 65, suggesting a potential reporting artifact (**Panels C and G in Figure 3; Panel C in Figure S5-S11; Figure S12)**. To assess whether this pattern could be explained by health insurance or Medicare coverage, we examined age distributions by insurance type (**Figure 4**). We also observe a notably strong increase in the number of individuals on Medicare in both the entire EHR dataset (0.08% to 4.6%) as well as individuals with menopause codes, although the sample size reporting health insurance type is notably small (i.e., only ∼1/10 response rate, N≈ 28,305) limiting our ability to determine whether Medicare coverage explains the spike in menopause EHR codes at 65 (**Figure 4**).

**Figure 3.**
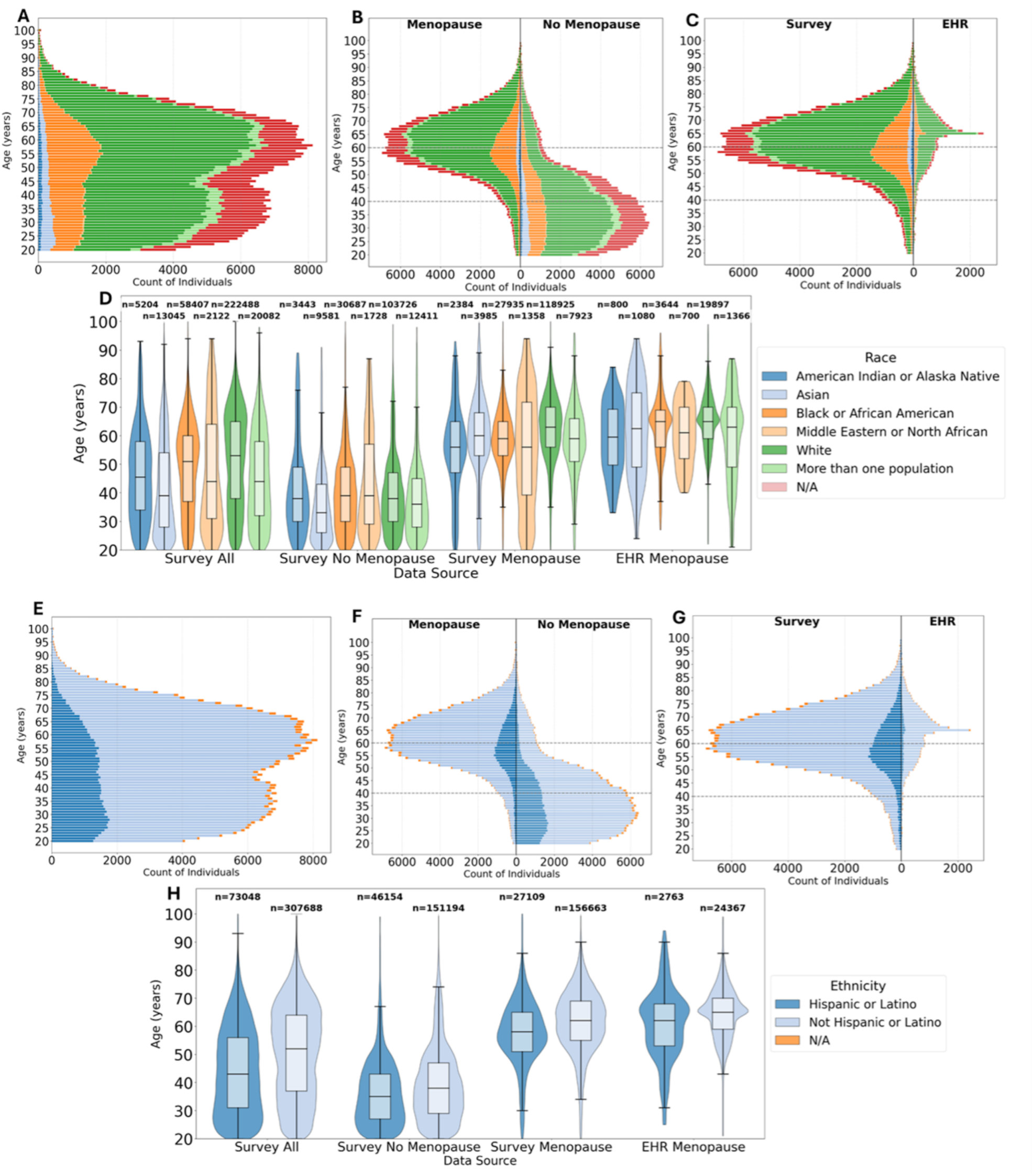
AoURP v8 age distributions by self-identified race and ethnicity. Histograms (1-year rounded bins) are shown by self-identified race **(A-C)** and ethnicity **(E-G)** for all female participants **(A, E)**, menopause survey response (answered “*yes none*” vs. “no” to “*Have your menstrual periods stopped permanently?*”) **(B, F)**, and menopause survey response (answered “*yes none*” to “*Have your menstrual periods stopped permanently?*”) vs. EHR menopause [SNOMED diagnostic code for *menopause present* (SNOMED 289903006)] **(C, G)**. Pirate plots of all females comparing survey and menopause general classifications are shown for self-identified race **(D)** and ethnicity **(H)**. Grey dashed lines in panels B, C, F and G indicate the approximate menopause transition age range (40-60 years). Sample sizes ≤20 are obscured and reported as “20” in accordance with AoURP Data Dissemination Policy. All other participant counts were rounded up to the nearest multiple of 5.

**Figure 4.**
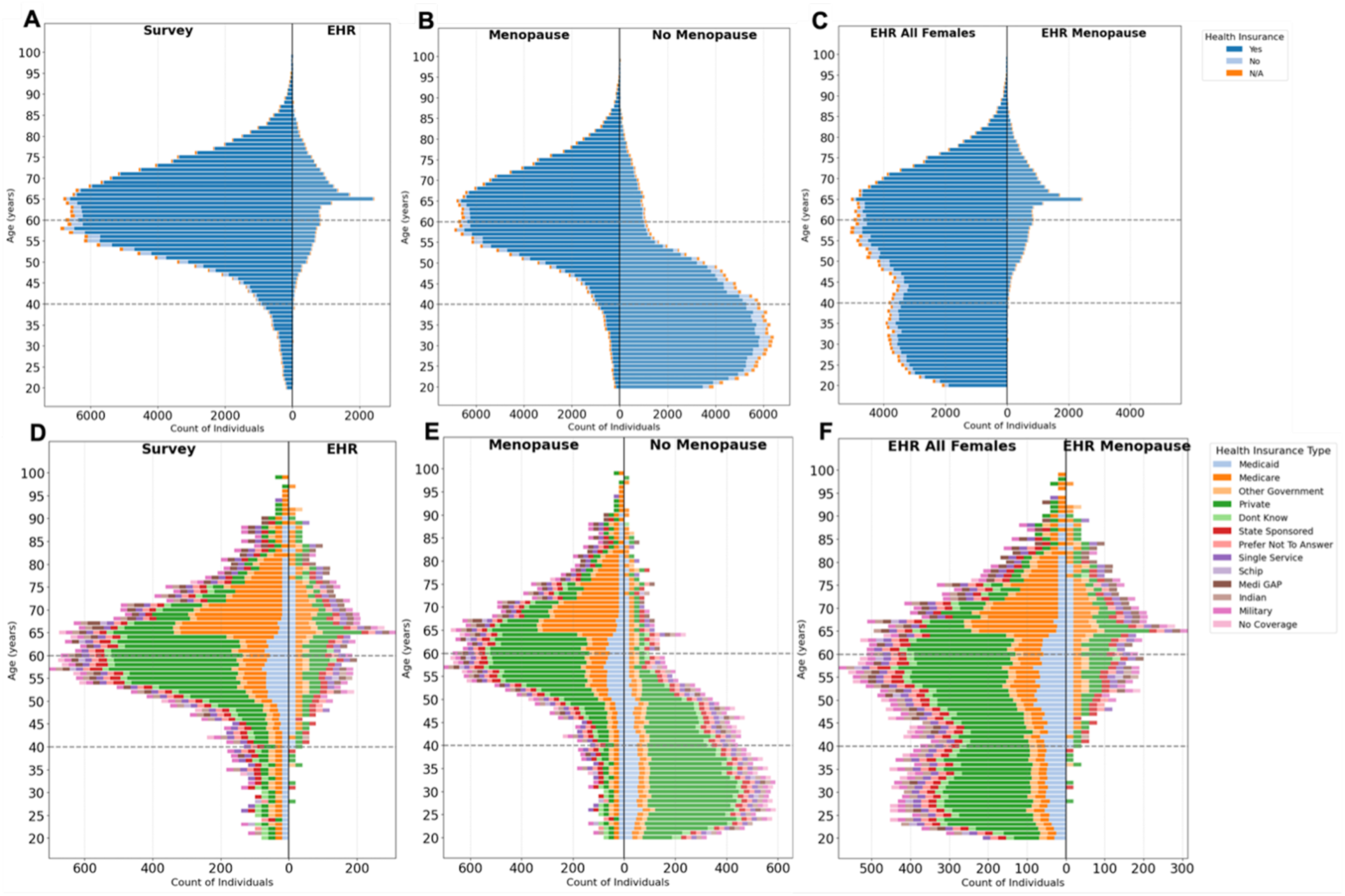
AoURP v8 age distributions by presence and type of health insurance. Histograms (1-year rounded bins) are shown for female participants for presence **(A-C)** and type **(D-F)** of health insurance: Menopause survey (answered “*yes none*” to the question “*Have your menstrual periods stopped permanently?*”) vs EHR [SNOMED diagnostic code for *menopause present* (SNOMED 289903006)] **(A, D)**, survey menopause status (answered “*yes none*” to “*Have your menstrual periods stopped permanently?*” question vs “*no*”) **(B, E)**, and general EHR data vs EHR menopause by health insurance type **(C, F)**. Grey dashed lines indicate the approximate menopause transition age range (40-60 years). Sample sizes ≤ 20 are obscured and reported as “20” in accordance with AoURP Data Dissemination Policy. All other participant counts were rounded up to the nearest multiple of 5.

### Age distributions by menopause and sociodemographic characteristics

Age distributions for “*yes*” versus “*no*” responses to the survey question “*Have your menstrual periods stopped permanently?*” matched expectations with menopause “*yes*” responses having older ages and “*no*” responses having younger ages for all demographic characteristics analyzed (**Panels B, D, F, and H in Figure 3; Panels B and D in Figure S5-S11**). As ages were documented at the time of the survey, not time of menopause, this consistency in general age distributions is important. Some female individuals reported not having gone through menopause at ages beyond biologically plausible ranges [e.g., N≈1,725 (4%) of females >70 years that answered the menopause survey question (N≈39,065)] (example in **Figure 3B**) indicating a possible response bias. The shape of the menopause survey age distribution did not differ substantially by most sociodemographic strata groups (**Figures 3, S5-S11**). Minor shifts were observed in some strata. For example, individuals who reported having gone through menopause and who self-reported White race (**Figure 3D, S5D**), self-reported non-Hispanic or Latino ethnicity (**Figure 3H, S5H**), or the European-like genetic ancestry group (**Figure S6-S7**) were slightly older than those of other race, ethnicity, and ancestry groups, respectively. However, these differences match those seen in the overall female AoURP sample suggesting any differences between strata are likely driven by underlying age distributions for the overall sample rather than menopause-specific effects (**Panel D and H in Figure 3, Panel D and H in Figure S5, Panel D in Figure S6-S7**). Supporting this, the female European-like genetic ancestry group sample has N≈139,595 individuals (∼56% of total females with genetic ancestry data) with median age of 56 years, whereas all other genetic ancestry groups had smaller sample sizes and younger median ages (African-like=50, Admixed American-like=43, East Asian-like=40, Middle Eastern-like=40, and South Asian-like=36 years) (**Table 1, S1**). Similarly, self-reported White female participants, most of whom also reported non-Hispanic or Latino ethnicity, had larger overall sample sizes and slightly older median ages than other race and ethnicity groups, respectively (**Table 1-2, S1-S2**).

## DISCUSSION

Here, we present a comprehensive characterization of menopause descriptors within the AoURP cohort. We computed descriptive summary statistics of menopause EHR diagnostic codes, menopause responses to women’s health questions (from a subsection of the AoURP “Overall Health” survey), and intersections with genomic data. Understanding how menopause data in AoURP are documented and represented informs future studies of the relationship between menopause status and health outcomes, including those integrating genomic data. Our findings demonstrate the complexity of identifying and describing menopause status in a large, sociodemographically diverse cohort using survey and EHR data, including evidence of response bias in the EHR. Moreover, our study highlights opportunities to improve systematic documentation of menopause status in the EHR and in the survey data. A key limitation of the AoURP dataset for menopause research is the absence of age at natural menopause in either the survey or EHR, an essential variable that should be captured in addition to menopause status.

Self-reported survey responses captured substantially more (∼7x) menopause observations than EHR diagnostic codes alone. The large difference between menopause counts in the EHR and survey aligns with prior research showing that EHRs often lack complete documentation of menopause status, even among individuals experiencing menopause symptoms or undergoing treatment^7–9^. Within AoURP, underreporting of menopause in the EHR does not appear to differ by menopause type as recorded in the survey (e.g., natural vs. surgical). Although identifying contributors to underreporting of menopause status in the AoURP EHR was not the focus of our study, underreporting may be due to factors such as inconsistent documentation practices, healthcare visits for conditions unrelated to menopause, and a lack of formal menopause diagnoses in the EHR unless linked to medication therapy or procedures for insurance and billing reasons, as previously described^7–9^. The diversity of menopause experiences observed is important for understanding downstream health risks of natural, surgical, and medication-induced menopause in future studies, as menopause status has implications for CVM health, among other health outcomes^4,6^. When surgeries occur prior to menopause, individuals may experience menopause differently or may not identify as menopausal, despite cessation of menstrual cycles. Underreporting of menopause in the EHR should be carefully considered when using EHR-derived menopause data for future AoURP studies. Conversely, response rates for the survey menopause question were substantially higher and thus would provide greater statistical power for future menopause focused research investigations. Our findings with the survey data align with prior work showing that self-reported menopause descriptors from surveys can be more comprehensively documented compared to EHR records and provide information on nuanced individual menopause experiences^7–9^. We also highlight incomplete or no capture of common menopause symptoms (e.g., specific symptoms: hot flashes, non-specific symptoms: sleep disturbances, mood alterations, joint aches and pains) in the both EHR and survey data. This finding is not only limited to the AoURP dataset, but common across EHR datasets in general due to lack of adequate diagnostic coding options for common menopause symptoms (e.g., vasomotor symptoms)^7–9^.

Additionally, sample size differences across sociodemographic strata, including race and genetic ancestry in the overall sample influence observed distributions of menopause information in surveys and EHRs, as we find here. While we illustrate general patterns, small sample sizes in some groups warrant caution in interpretation, especially when evaluating age distributions or drawing comparisons across strata. Ultimately, any differences or trends observed in menopause characteristics across the contexts investigated here (e.g., ethnicity, race) are descriptive and likely reflect the general demographic makeup of the female AoURP cohort rather than specific aspects of menopause. Importantly, sociodemographic representation in the AoURP cohort is also likely influenced by additional factors (e.g., geographic location of clinical sites, healthcare access barriers, willingness to participate in biomedical research).

We provide summary statistics of menopause descriptors across data types highlighting the opportunity for integrated, multimodal analyses within AoURP that go beyond what single data sources can provide. Notably, sample sizes are considerably smaller when intersecting with EHR data. EHR and survey datas intersected with genomic data provide a strong foundation for future genomic and precision medicine research, where integrating menopause status with genomic variation can advance understanding of genomic and environmental influences on menopause timing, symptoms, and other health outcomes^4,6^. Moving forward, researchers can leverage this multimodal AoURP resource to design socio-demographically informed studies aimed at understanding the complex relationships between menopause and genomics.

## CONCLUSIONS

Our comprehensive summary of menopause descriptors in the AoURP serves as a foundation for researchers designing menopause studies with AoURP data. This study provides insight into available menopause variables, the sample sizes for sociodemographic strata and subgroups, and the overlap across EHR, survey, and genomic data types. Future studies can build on this work to investigate clinical outcomes associated with menopause (e.g., CVM health), potentially in combination with genomic data. By highlighting where menopause information is or is not well-documented and well-represented in the AoURP dataset, our findings can guide researchers in designing sufficiently powered, socio-demographically representative studies for future investigations. More broadly, these findings contribute to the growing efforts to improve data accessibility, quality, and interoperability for menopause research in large, comprehensive national cohorts representing the diverse characteristics of the US. These findings also help inform improvements in data documentation and collection of menopause status across EHR, survey, and genomic data types as well as all sociodemographic strata within AoURP.

## POTENTIAL CLINICAL VALUE

This work has potential clinical value by showing that menopause data is commonly underascertained in structured EHR data relative to self-reported survey data, which may affect identification of menopause-related data in both research and clinical settings. Better characterization of menopause status by including age of menopause instead of time of menopause data observation in the AoURP EHR and survey data as well as other studies could improve menopause data studies, support more accurate demographic stratification for analyses of menopause associations with other midlife health outcomes, and guide future studies of genetic and sociodemographic influences on menopause-related health trajectories.

These findings may ultimately contribute to improved ascertainment of menopause data in large-scale biobank studies and EHR data in general as well as inform sampling strategies for future observational and translational studies with menopause data to ultimately improve health equity for all.

## Supporting information

Supplemental Digital Content 1.pdf

Supplemental Digital Content 2.xlsx

## Data Availability

The original data used for this analysis cannot be shared publicly in accordance with All of Us Research Program data sharing policies, but can be accessed through the All of Us Research Program Researcher Workbench for registered and approved users.

## ACKNOWLEDGEMENTS

We gratefully acknowledge the All of Us participants for their contributions, without whom this research would not have been possible. We also thank the National Institutes of Health’s All of Us Research Program for making available the participant data examined in this study. We also acknowledge the support and guidance from the Ludeman Family Center for Women’s Health Research.

## AUTHOR APPROVAL

All authors have seen and approved the manuscript.

## DECLARATIONS

This manuscript has not been presented in any format at a national meeting.

## DATA AVAILABILITY STATEMENT

The original data used for this analysis cannot be shared publicly in accordance with All of Us Research Program data sharing policies.

## SUPPLEMENTAL DIGITAL CONTENT

1. **Supplemental Digital Content 1 (Supplemental Digital Content 1.pdf)** – A PDF document of supplemental results tables and figures.
2. **Supplemental Digital Content 2 (Supplemental Digital Content 2.xlsx)** – A Microsoft Excel file (.xlsx) with supplemental results tables.

## REFERENCES

1. Harlow SD, Gass M, Hall JE, et al. Executive summary of the Stages of Reproductive Aging Workshop + 10: addressing the unfinished agenda of staging reproductive aging. Menopause N Y N. 2012;19(4):387–395. doi:10.1097/gme.0b013e31824d8f40

2. Gold EB. The timing of the age at which natural menopause occurs. Obstet Gynecol Clin North Am. 2011;38(3):425–440. doi:10.1016/j.ogc.2011.05.002

3. Schoenaker DAJM, Jackson CA, Rowlands JV, Mishra GD. Socioeconomic position, lifestyle factors and age at natural menopause: a systematic review and meta-analyses of studies across six continents. Int J Epidemiol. 2014;43(5):1542–1562. doi:10.1093/ije/dyu094

4. Anagnostis P, Stevenson JC. Cardiovascular health and the menopause, metabolic health. Best Pract Res Clin Endocrinol Metab. 2024;38(1):101781. doi:10.1016/j.beem.2023.101781

5. Management of osteoporosis in postmenopausal women: the 2021 position statement of The North American Menopause Society. Menopause N Y N. 2021;28(9):973–997. doi:10.1097/GME.0000000000001831

6. El Khoudary SR, Aggarwal B, Beckie TM, et al. Menopause Transition and Cardiovascular Disease Risk: Implications for Timing of Early Prevention: A Scientific Statement From the American Heart Association. Circulation. 2020;142(25):e506–e532. doi:10.1161/CIR.0000000000000912

7. Eyre H, Alba PR, Gibson CJ, et al. Bridging information gaps in menopause status classification through natural language processing. JAMIA Open. 2024;7(1):ooae013. doi:10.1093/jamiaopen/ooae013

8. Sussman M, Trocio J, Best C, et al. Prevalence of menopausal symptoms among mid-life women: findings from electronic medical records. BMC Womens Health. 2015;15:58. doi:10.1186/s12905-015-0217-y

9. Bevry ML, Stogdill ER, Lea CM, et al. Addressing menopause symptoms in the primary care setting: opportunity to bridge care delivery gaps. Menopause N Y N. 2024;31(12):1044–1048. doi:10.1097/GME.0000000000002439

10. Bycroft C, Freeman C, Petkova D, et al. The UK Biobank resource with deep phenotyping and genomic data. Nature. 2018;562(7726):203–209. doi:10.1038/s41586-018-0579-z

11. Santoro N. The Study of Women’s Health Across the Nation (SWAN). Obstet Gynecol Clin North Am. 2011;38(3):xvii-xix. doi:10.1016/j.ogc.2011.07.002

12. El Khoudary SR, Greendale G, Crawford SL, et al. The menopause transition and women’s health at midlife: a progress report from the Study of Women’s Health Across the Nation (SWAN). Menopause. 2019;26(10):1213–1227. doi:10.1097/GME.0000000000001424

13. All of Us Research Program Genomics Investigators. Genomic data in the All of Us Research Program. Nature. 2024;627(8003):340–346. doi:10.1038/s41586-023-06957-x

14. Baskir R, Lee M, McMaster SJ, et al. Research for all: building a diverse researcher community for the All of Us Research Program. J Am Med Inform Assoc JAMIA. 2025;32(1):38–50. doi:10.1093/jamia/ocae270

15. Kochersberger A, Coakley A, Millheiser L, et al. The association of race, ethnicity, and socioeconomic status on the severity of menopause symptoms: a study of 68,864 women. Menopause N Y N. 2024;31(6):476–483. doi:10.1097/GME.0000000000002349

16. Cortés YI, Marginean V. Key factors in menopause health disparities and inequities: Beyond race and ethnicity. Curr Opin Endocr Metab Res. 2022;26:100389. doi:10.1016/j.coemr.2022.100389

17. Chamberlin DD, Boyce RF. SEQUEL: A structured English query language. In: Proceedings of the 1976 ACM SIGFIDET (Now SIGMOD) Workshop on Data Description, Access and Control - FIDET ’76. ACM Press; 1976:249–264. doi:10.1145/800296.811515

18. R Core Team. R: A Language and Environment for Statistical Computing, version 4.5.0. R Foundation for Statistical Computing, Vienna, Austria. 2025. https://www.R-project.org/

19. Python Software Foundation. Python Language Reference, version 3.10.16. Published online 2025. https://www.python.org

20. Wickham H. tidyverse: Easily Install and Load the “Tidyverse.” Published online September 9, 2016. doi:10.32614/cran.package.tidyverse

21. Wickham H, Bryan J. bigrquery: An Interface to Google’s “BigQuery” “API.” Published online January 13, 2015. doi:10.32614/cran.package.bigrquery

22. Wickham H, Pedersen TL, Seidel D. scales: Scale Functions for Visualization. Published online September 22, 2011. doi:10.32614/cran.package.scales

23. Dave A, Patel DJ, Shrivastava D, Chaudhari K, Manchanda R. Considerations in Premature Menopause: A Review. Cureus. 2024;16(9):e69744. doi:10.7759/cureus.69744

24. Paramsothy P, Harlow SD, Nan B, et al. Duration of the menopausal transition is longer in women with young age at onset: the multiethnic Study of Women’s Health Across the Nation. Menopause. 2017;24(2):142–149. doi:10.1097/GME.0000000000000736

25. Luborsky JL, Meyer P, Sowers MF, Gold EB, Santoro N. Premature menopause in a multi-ethnic population study of the menopause transition. Hum Reprod. 2003;18(1):199–206. doi:10.1093/humrep/deg005

26. All of Us Research Program Support Page. All of Us Survey Data Codebooks with Privacy Rules. March 25, 2022. Accessed February 27, 2026. https://docs.google.com/spreadsheets/d/15b4KEchI9fUcaG42DVEf9ikl6UpVtdM0GZrjFMKMrR4/edit?pli=1&gid=1558751632#gid=1558751632

27. All of Us Research Program Support Page. Overall Health: Baseline Survey Details and Resources. 2024. Accessed February 27, 2026. https://support.researchallofus.org/hc/en-us/articles/6085831989524-Overall-Health-Baseline-Survey-Details-and-Resources

