## Supplemental Digital Content 1.pdf for "Menopause in the All of Us Research Program: A Descriptive Summary of Electronic Health Record and Survey Response across Sociodemographic Characteristics"

### Table S1. AoURP v7 sample sizes and age distributions for sociodemographic and menopause variables for female participants.

| Sociodemographic strata group | N (% of group) | Age (years) summary[a] | | | |
|---|---|---|---|---|---|
| | | Q1 | Median | Q3 | IQR |
| **Full AoURP (sex = female)** | 249565 (100%) | 35 | 50 | 63 | 28 |
| **Age quartile** | | | | | |
| Q1 (19-41 years) | 88815 (35.8%) | 25 | 31 | 36 | 11 |
| Q2 (42-57 years) | 70530 (28.4%) | 46 | 50 | 54 | 8 |
| Q3 (58-69 years) | 57565 (23.2%) | 60 | 63 | 66 | 6 |
| Q4 (≥70 years) | 31075 (12.5%) | 71 | 74 | 78 | 7 |
| **Age (menopause transition)** | | | | | |
| <40 years | 82500 (33.1%) | 25 | 30 | 35 | 10 |
| 40-60 years | 93750 (37.6%) | 46 | 51 | 56 | 10 |
| >60 years | 73325 (29.4%) | 64 | 68 | 73 | 9 |
| **Education level (EL)** | | | | | |
| EL1 (never attend) | 365 (0.1%) | 50 | 61 | 70 | 20 |
| EL2 (grade 1-4) | 2235 (0.9%) | 48 | 58 | 67 | 19 |
| EL3 (grade 5-8) | 5535 (2.2%) | 43 | 53 | 63 | 20 |
| EL4 (grade 9-11) | 13170 (5.3%) | 35 | 48 | 57 | 22 |
| EL5 (grade 12, GED) | 43595 (17.5%) | 32 | 48 | 60 | 28 |
| EL6 (college 1-3 years) | 67500 (27.1%) | 34 | 50 | 62 | 28 |
| EL7 (college graduate) | 59540 (23.9%) | 32 | 49 | 62 | 30 |
| EL8 (advanced degree) | 53105 (21.3%) | 38 | 53 | 66 | 28 |
| prefer not to answer, skip | 4465 (1.8%) | 39 | 52 | 62 | 23 |
| **Ethnicity** | | | | | |
| Hispanic or Latino | 49410 (19.8%) | 30 | 42 | 55 | 25 |
| not Hispanic or Latino | 193770 (77.6%) | 36 | 52 | 64 | 28 |
| prefer not to answer, skip | 6395 (2.6%) | 39 | 54 | 66 | 27 |
| **Income level (IL)** | | | | | |
| IL1 (<10k) | 31380 (12.6%) | 31 | 45 | 56 | 25 |
| IL2 (10k-25k) | 29505 (11.8%) | 35 | 51 | 63 | 28 |
| IL3 (25k-35k) | 19320 (7.8%) | 31 | 46 | 63 | 32 |
| IL4 (35k-50k) | 21210 (8.5%) | 32 | 48 | 64 | 32 |
| IL5 (50k-75k) | 27770 (11.1%) | 34 | 51 | 65 | 31 |
| IL6 (75k–100k) | 21085 (8.5%) | 36 | 53 | 65 | 29 |
| IL7 (100k–150k) | 24995 (10%) | 38 | 52 | 63 | 25 |
| IL8 (150k–200k) | 11615 (4.7%) | 38 | 51 | 62 | 24 |
| IL9 (>200k) | 14440 (5.8%) | 40 | 52 | 61 | 21 |
| prefer not to answer, skip | 47915 (19.2%) | 35 | 52 | 64 | 29 |
| **Race** | | | | | |
| Asian | 8535 (3.4%) | 27 | 38 | 54 | 27 |
| Black African American | 45280 (18.1%) | 36 | 50 | 60 | 24 |
| Middle East North Africa | 1290 (0.5%) | 29 | 39 | 56 | 27 |
| more than one population | 5195 (2.1%) | 27 | 36 | 52 | 25 |
| Native Hawaii Pacific Island | 255 (0.1%) | 30 | 43 | 54 | 24 |
| White | 139380 (55.8%) | 37 | 54 | 66 | 29 |
| prefer not to answer, skip | 49640 (19.9%) | 31 | 45 | 57 | 26 |
| **Ancestry (requires genetic data)** | | | | | |
| AFR-like | 31980 (22%) | 36 | 50 | 60 | 24 |
| AMR-like | 29570 (20.3%) | 30 | 43 | 56 | 26 |
| EAS-like | 3590 (2.5%) | 28 | 41 | 57 | 29 |
| EUR-like | 78250 (53.8%) | 39 | 56 | 67 | 28 |
| MID-like | 490 (0.3%) | 27 | 39 | 55 | 28 |
| SAS-like | 1695 (1.2%) | 27 | 37 | 51 | 24 |
| **Menopause** | | | | | |
| EHR [*premature* or *menopause present*][b] | 18280 [7.3% of all females (N=249565)] | 58 | 65 | 70 | 12 |
| Survey [yes for any reason][c] | 129255 (87.6%) [51.8% of all females (N=249565)] | 54 | 62 | 69 | 15 |

Abbreviations: AoURP, All of Us Research Program; EHR, electronic health record; EL, education level; IL, income level; IQR, interquartile range; N, number of participants; Q1, first quartile; Q3, third quartile. For education and income level, prefer not to answer or skip responses were combined. For ethnicity, none of these or prefer not to answer or skip responses were combined. For race, "*None Indicated*" or "*None of these*" or prefer not to answer or skip responses were combined. Participant counts ≤20 are reported as "N ≤ 20" in accordance with the AoURP Data Dissemination Policy. All other participant counts were rounded up to nearest 5. [a]Age summary statistics include Q1, median, Q3, and IQR, calculated from birth date and demographic basic survey date; age for menopause descriptors was calculated from birth date and descriptor earliest observation date in EHR or survey. [b]EHR menopause diagnostic codes: *premature menopause* (SNOMED 373717006) or *menopause present* (SNOMED 289903006). [c]Survey menopause: answered yes for any reason to "*Have your menstrual periods stopped permanently?*" question.

**Table S2. Sample sizes of AoURP v7 female participants by self-reported race and ethnicity.**

| Race | Ethnicity (N) | | |
|---|---|---|---|
| | Hispanic or Latino (% of row total) | Not Hispanic or Latino (% of row total) | Total (% of column total) |
| Asian | 280 (3.3%) | 8260 (96.7%) | 8540 (3.5%) |
| Black African American | 890 (2%) | 44395 (98%) | 45285 (18.6%) |
| Middle East North Africa | 105 (8.1%) | 1190 (91.9%) | 1295 (0.5%) |
| more than one population | 630 (12.1%) | 4565 (87.9%) | 5195 (2.1%) |
| Native Hawaii Pacific Island | 35 (13.5%) | 225 (86.5%) | 260 (0.1%) |
| White | 4235 (3%) | 135145 (97%) | 139380 (57.3%) |
| Skip | 43250 (99.95%) | N ≤ 20 (0.05%) | 43270 (17.8%) |

Abbreviations: N, number of participants. Participant counts ≤20 are reported as "N ≤ 20" in accordance with the AoURP Data Dissemination Policy. All other participant counts were rounded up to nearest 5. Sample size of participants that skipped both race and ethnicity questions≈6395.

**Table S3. AoURP v7 survey and EHR diagnostic descriptors of menopause for female participants.**

| Survey question | Survey response | N (%)[a] |
|---|---|---|
| **Have your menstrual periods stopped permanently?** | **Total (sum of menstrual stop responses)** | **248820 (100%)** |
| | Yes none | 126740 (50.9%) |
| | Yes but hormone | 2515 (1.0%) |
| | **Total yes (sum of rounded yes responses)** | **129255 (51.9%)** |
| | Periods have not stopped | 99955 (40.2%) |
| | Not sure | 6085 (2.4%) |
| | Prefer not answer | 3670 (1.5%) |
| | Skip | 9855 (4.0%) |
| **— Stopped reason** | **Total (sum of rounded stop reason responses)** | **131670 (100%)** |
| | Natural menopause | 73610 (55.9%) |
| | Surgery | 39795 (30.2%) |
| | Other | 4650 (3.5%) |
| | Medication therapy | 4775 (3.6%) |
| | Endometrial ablation | 3475 (2.6%) |
| | Skip | 1965 (1.5%) |
| | Not sure | 2505 (1.9%) |
| | Prefer not answer | 895 (0.7%) |
| **Hysterectomy history** | **Total (sum of rounded hysterectomy responses)** | **141420 (100%)** |
| | No | 91550 (64.7%) |
| | Yes | 45475 (32.2%) |
| | Skip | 2005 (1.4%) |
| | Prefer not answer | 1635 (1.2%) |
| | Not sure | 755 (0.5%) |
| **Ovary removed history** | **Total (sum of rounded ovary removed responses)** | 141425 (100%) |
| | Yes both | **22115 (15.6%)** |
| | Yes partial (sectioned) | 9555 (6.8%) |
| | Yes unsure | 785 (0.6%) |
| | **Total yes (sum of rounded yes responses)** | **32455 (22.9%)** |
| | No | 103135 (72.9%) |
| | Skip | 2145 (1.5%) |
| | Not sure | 2190 (1.5%) |
| | Prefer not answer | 1500 (1.1%) |
| **EHR condition** | **SNOMED Code** | **N (%)[a]** |
| **Menopause** | **Total menopause [least one Menopause present (289903006) or Premature menopause (373717006) code]** | **18280 (100%)** |
| | Menopause present (289903006)[b] | 17675 (96.7% of N=18280) |
| | - Individuals with 1 code | 7585 (42.9% of N=17675) |
| | - Individuals with 2 codes | 4575 (25.9% of N=17675) |
| | - Individuals with 3 codes | 1570 (8.9% of N=17675) |
| | - Individuals with 4 codes | 1520 (8.6% of N=17675) |
| | - Individuals with 5 codes | 550 (3.1% of N=17675) |
| | - Individuals with > 1 code | 10090 (57.1% of N=17675) |
| | - Individuals with > 5 code | 1880 (10.6% of N=17675) |
| | Premature menopause (373717006)[c] | 870 (4.8% of N=18280) |
| **Ovarian failure** | **Total ovarian failure (sum of rounded ovarian failure EHR diagnostic codes)** | **3665 (100%)** |
| | Primary ovarian failure (65846009) | 3625 (98.9%) |
| | Premature ovarian failure (237788002) | N ≤ 20 (≤0.5%) |
| | Menopause ovarian failure (237138004) | N ≤ 20 (≤0.5%) |
| **Vasomotor symptoms** | Abnormal vasomotor function (70670009) | N ≤ 20 |

Abbreviations: EHR, electronic health record; N, number of participants; SNOMED, Systematized Nomenclature of Medicine. Survey data include self-reported menstrual history, menopause reason, and surgical history. EHR data are defined by menopause SNOMED condition codes. [1]Participant counts ≤20 are reported as "N ≤ 20" in accordance with AoURP Data Dissemination Policy; All other participant counts were rounded up to nearest 5.

[a]% is proportion of total number in each survey question or EHR condition category.

[b]Count also includes individuals that have premature menopause (373717006) code.

[c]Count also includes individuals that have menopause present (289903006) code.

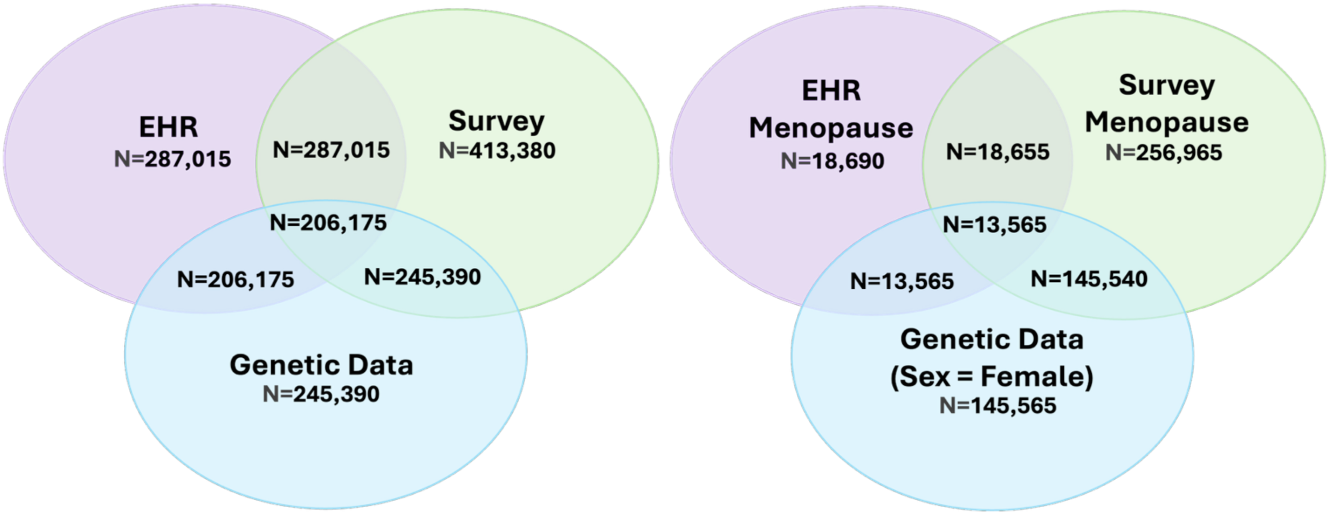


**Figure S1. Intersections of EHR, survey, and genomic data for all AoURP v7 participants.** Venn diagram illustrating the intersection sample sizes of participants with EHR, survey, and genomic data modalities for all participants **(left)** and menopause questions/indications and female participants **(right)**. The EHR Menopause group includes unique participants with at least one SNOMED diagnostic code for *menopause present* (289903006) or *premature menopause* (373717006). The Survey Menopause group includes participants who responded with any answer to the "*Have your menstrual periods stopped permanently?*" question. The Genomic Data (Sex = Female) group is female sex individuals with srWGS data. All participant counts were rounded up to nearest value of 5.

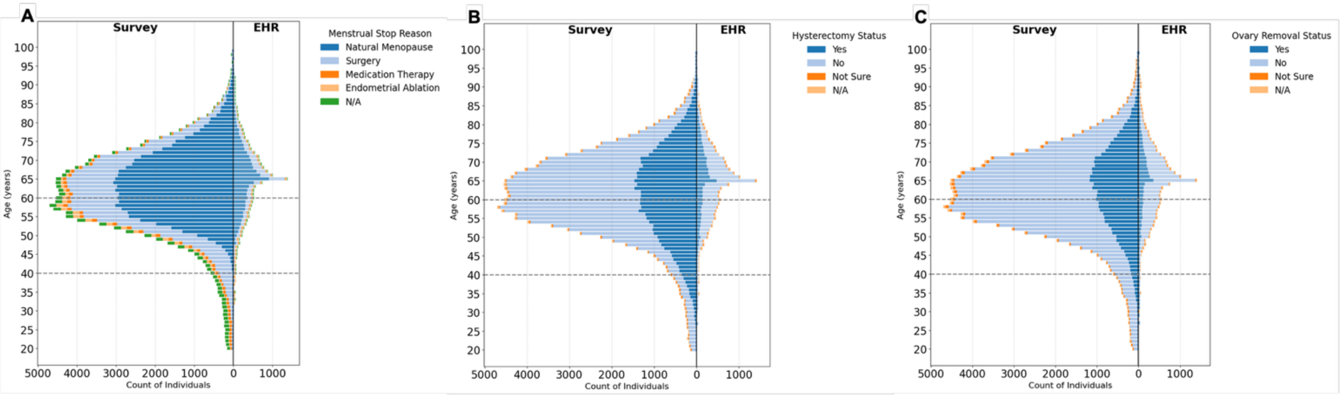


**Figure S2. AoURP v7 age distributions in the survey and EHR for survey menopause classifications.** Histograms with 1-year rounded bins for participants with female sex at birth for survey (answered "*Have your menstrual periods stopped permanently?*" question) **(left panels)** vs. EHR menopause status [SNOMED diagnostic code for *menopause present* (289903006)] **(right panels)** for **(A)** survey reason for menopause (*"menstrual stop reason"*), **(B)** survey "*hysterectomy history*" status, and **(C)** survey "*ovary removal history*" status. Histograms with 1-year rounded bins for survey (left panels) vs. Grey dashed lines indicate approximate menopause transition age range (40 to 60 years). Sample sizes ≤ 20 are obscured and reported as "20" in accordance with AoURP Data Dissemination Policy. All other participant counts were rounded up to nearest 5.

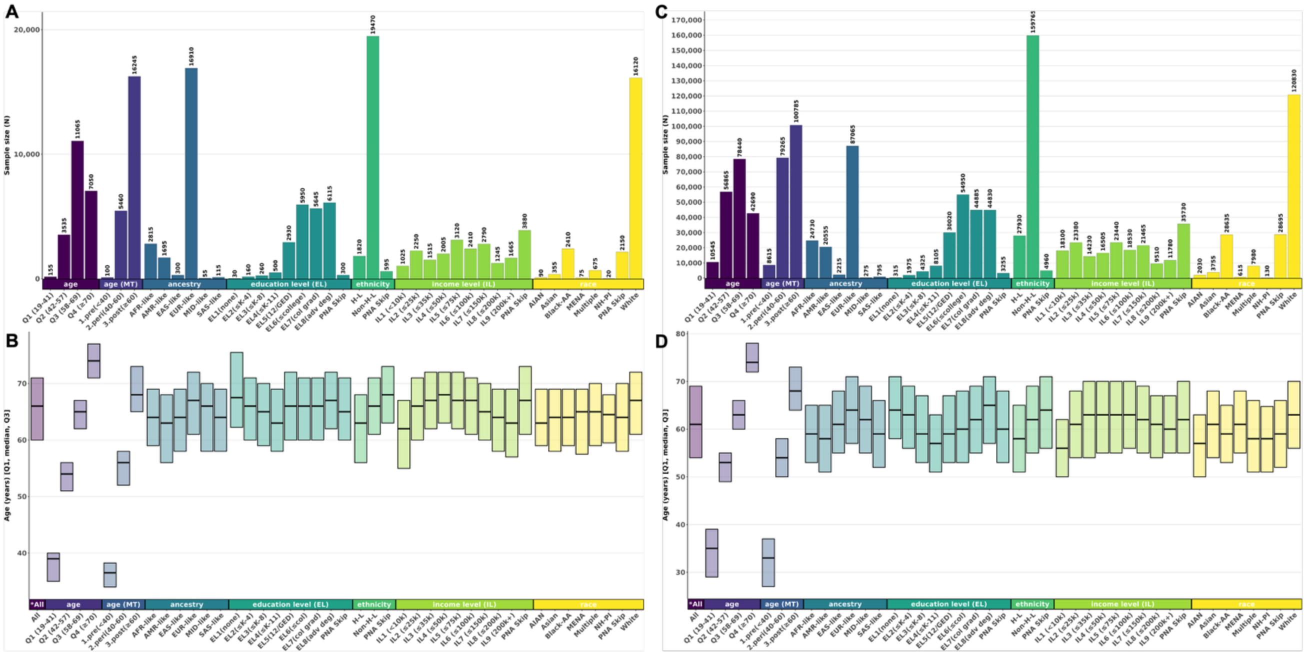


**Figure S3. AoURP v8 sample sizes and age distributions for female participants.** Bar charts for sample sizes for the three-way intersection across all three data sources, EHR, survey responses, and genomic data, by sociodemographic characteristics are shown in **(A)** and all survey menopause responses in **(C);** inter-quartile range and median for the three-way intersection are shown in **(B)** and all survey responses in **(D)**. The EHR group includes participants with at least one SNOMED diagnostic code for *menopause present* (289903006) or *premature menopause* (373717006). The survey group includes participants who responded "*yes none*" or "*yes but hormone*" to the question "*Have your menstrual periods stopped permanently?*" The genomic data group includes all female participants with available srWGS data. The survey responses include female participants who responded "*yes none*" or "*yes but hormone*" to the question "*Have your menstrual periods stopped permanently?*" Sociodemographic strata and groups (colored and ordered left to right) include: (1) *overall* (dark purple): all participants (no sociodemographic stratification) included for each data source ("*All"); (2) *age quartile* (purple): Q1 (19–41), Q2 (42–57), Q3 (58–69), Q4 (≥70); (3) *age (menopause transition age group)* (purple-blue): 1.age<40), 2.age 40–60, 3.age>60); (4) *education level (EL)* (dark blue): EL1 (none), EL2 (≤K–4), EL3 (≤K–8), EL4 (≤K–11), EL5 (12/GED), EL6 (≤college), EL7 (college graduate), EL8 (advanced degree), "*prefer not to answer*" or Skip (PNA Skip); (5) *ethnicity* (light blue): Hispanic or Latino (H-L), not Hispanic or Latino (Non-H-L), none of these or PNA or Skip (PNA Skip); (6) *income level (IL)* (green): IL1 (<10k), IL2 (≤25k), IL3 (≤35k), IL4 (≤50k), IL5 (≤75k), IL6 (≤100k), IL7 (≤150k), IL8 (≤200k), IL9 (200k+), PNA or Skip (PNA Skip); (7) *race* (green-yellow): American Indian Alaska Native (AIAN), Asian, Black or African American (Black-AA), Middle Eastern North African (MENA), more than one population (Multiple), Native Hawaiian or Other Pacific Islander (NH-PI), "*None Indicated*" or "*None of these*" or PNA or Skip (PNA Skip), White. Sample sizes ≤20 are obscured and reported as "20" in accordance with the AoURP Data Dissemination Policy. All other participant counts were rounded up to nearest 5.

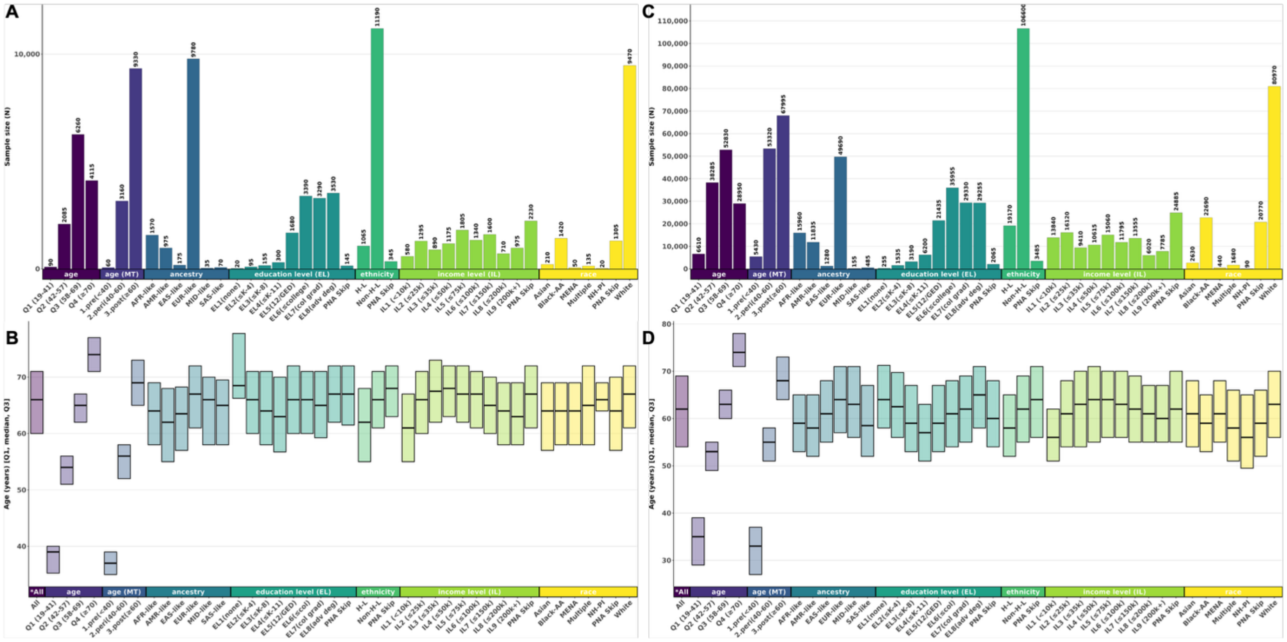


**Figure S4. AoURP v7 sample sizes and age distribution for female participants.** Bar charts for sample sizes for the three-way intersection across all three data sources, EHR, survey responses, and genomic data, by sociodemographic characteristics are shown in **(A)** and all survey menopause responses in **(C);** inter-quartile range and median for the three-way intersection are shown in **(B)** and all survey responses in **(D)**. The EHR group includes participants with at least one SNOMED diagnostic code for *menopause present* (289903006) or *premature menopause* (373717006). The survey group includes participants who responded "*yes none*" or "*yes but hormone*" to the question "*Have your menstrual periods stopped permanently?*" The genomic data group includes all female participants with available srWGS data. The survey responses include female participants who responded "*yes none*" or "*yes but hormone*" to the question "*Have your menstrual periods stopped permanently?*" Sociodemographic strata and groups (colored and ordered left to right) include: (1) *overall* (dark purple): all participants (no sociodemographic stratification) included for each data source ("*All"); (2) *age quartile* (purple): Q1 (19–41), Q2 (42–57), Q3 (58–69), Q4 (≥70); (3) *age (menopause transition age group)* (purple-blue): 1.age<40), 2.age 40–60, 3.age>60); (4) *education level (EL)* (dark blue): EL1 (none), EL2 (≤K–4), EL3 (≤K–8), EL4 (≤K–11), EL5 (12/GED), EL6 (≤college), EL7 (college graduate), EL8 (advanced degree), "*prefer not to answer*" or Skip (PNA Skip); (5) *ethnicity* (light blue): Hispanic or Latino (H-L), not Hispanic or Latino (Non-H-L), none of these or PNA or Skip (PNA Skip); (6) *income level (IL)* (green): IL1 (<10k), IL2 (≤25k), IL3 (≤35k), IL4 (≤50k), IL5 (≤75k), IL6 (≤100k), IL7 (≤150k), IL8 (≤200k), IL9 (200k+), PNA or Skip (PNA Skip); (7) *race* (green-yellow): American Indian Alaska Native (AIAN), Asian, Black or African American (Black-AA), Middle Eastern North African (MENA), more than one population (Multiple), Native Hawaiian or Other Pacific Islander (NH-PI), "*None Indicated*" or "*None of these*" or PNA or Skip (PNA Skip), White. Sample sizes ≤20 are obscured and reported as "20" in accordance with the AoURP Data Dissemination Policy. All other participant counts were rounded up to nearest 5.

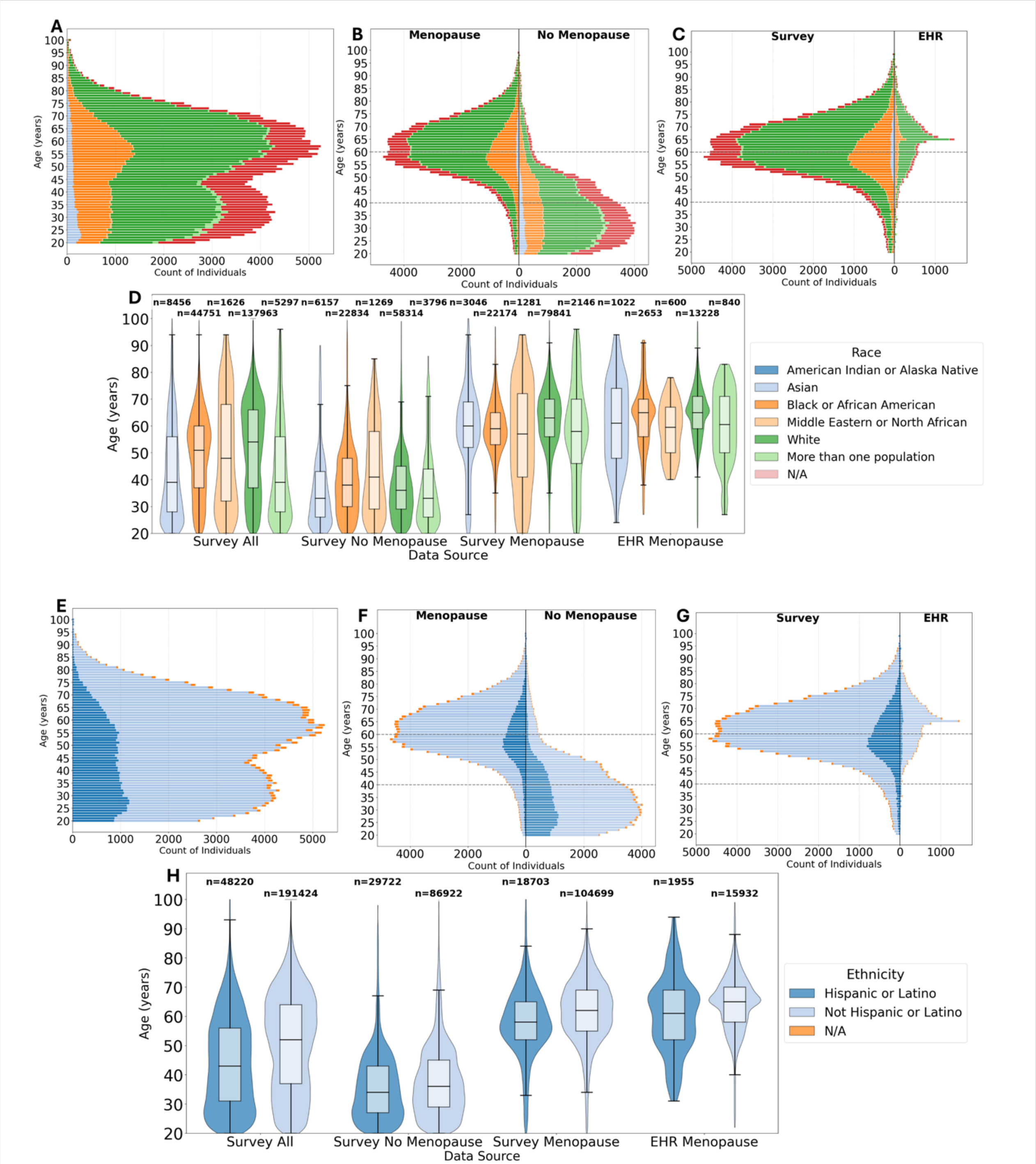


**Figure S5. AoURP v7 age distributions by self-identified race and ethnicity.** Histograms with 1-year rounded bins for self-identified race **(A-C)** and ethnicity **(E-G)** for all female participants **(A & E)**, menopause survey response (answered “*yes none*” vs. “no” to “*Have your menstrual periods stopped permanently?*”) **(B & F)**, and menopause survey response (answered “*yes none*” to “*Have your menstrual periods stopped permanently?*”) vs. EHR menopause [SNOMED diagnostic code for *menopause present* (289903006)] **(C & G)**. Pirate plots of all females comparing all survey and menopause general classifications are presented for self-identified race **(D)** and ethnicity **(H)**. Grey dashed lines in panel B, C, F, and G indicate approximate menopause transition age range (40 to 60 years). Sample sizes ≤ 20 are obscured and reported as “20” in accordance with AoURP Data Dissemination Policy. All other participant counts were rounded up to nearest 5.

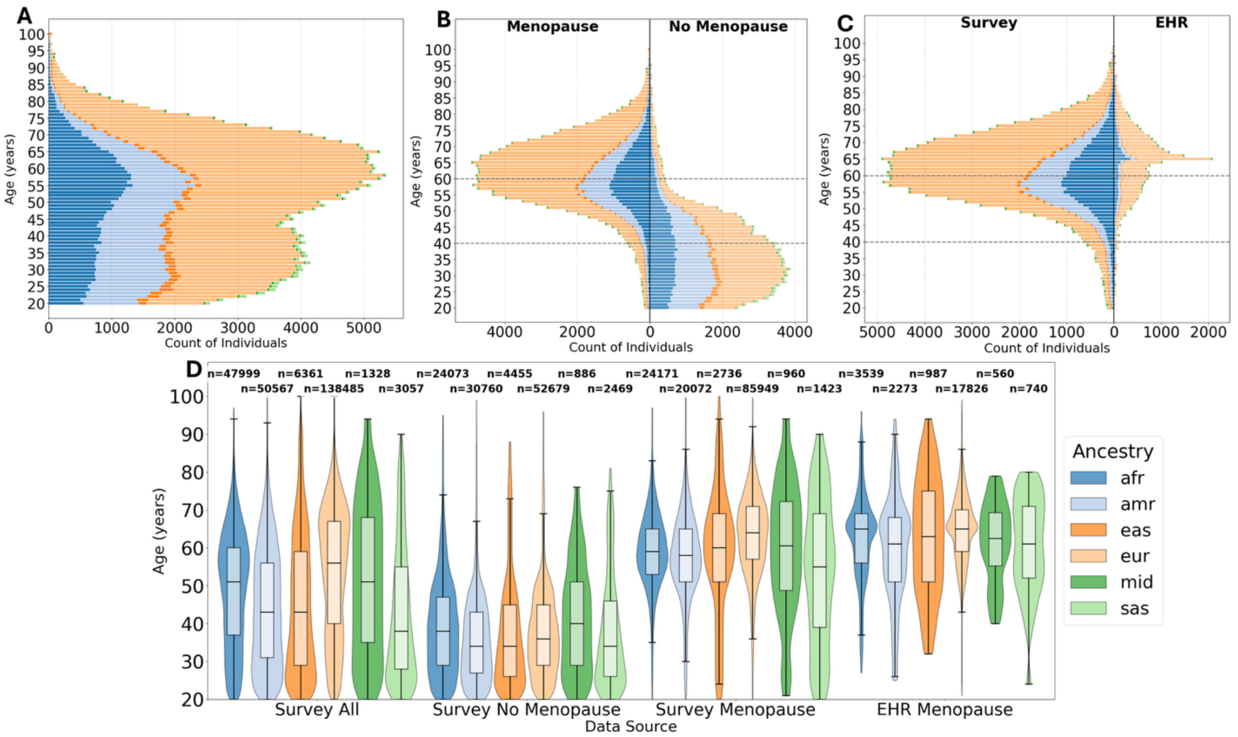


**Figure S6. AoURP v8 age distributions by genetic ancestry similarity group.** Histograms with 1-year rounded bins for all female participants **(A)**, menopause survey response (answered “*yes none*” vs. “no” to “*Have your menstrual periods stopped permanently?*”) **(B)**, and menopause survey response (answered “*yes none*” to “*Have your menstrual periods stopped permanently?*”) vs. EHR menopause [SNOMED diagnostic code for *menopause present* (289903006)] **(C)**. Pirate plots of all females comparing survey and menopause general classifications **(D)**. Grey dashed lines in panel B and C indicate approximate menopause transition age range (40 to 60 years). Sample sizes ≤ 20 are obscured and reported as “20” in accordance with AoURP Data Dissemination Policy. All other participant counts were rounded up to nearest 5.

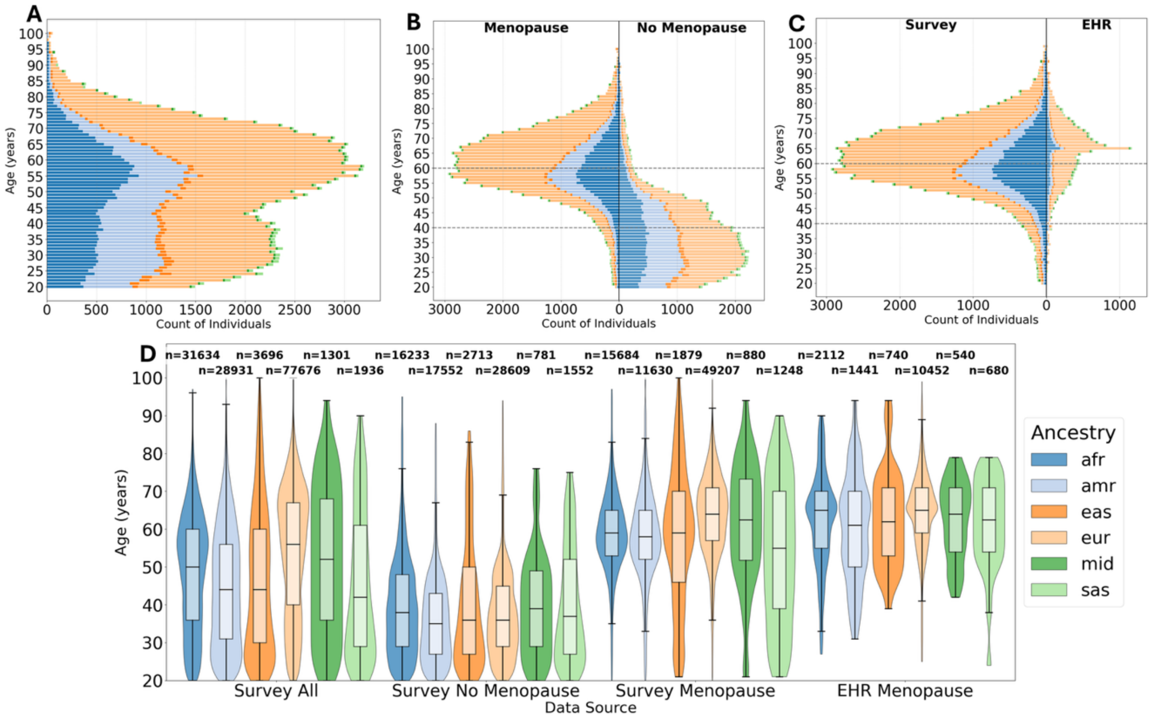


**Figure S7. AoURP v7 age distributions by genetic ancestry similarity group.** Histograms with 1-year rounded bins for all female participants **(A)**, menopause survey response (answered "*yes none*" vs. "no" to "*Have your menstrual periods stopped permanently?*") **(B)**, and menopause survey response (answered "*yes none*" to "*Have your menstrual periods stopped permanently?*") vs. EHR menopause [SNOMED diagnostic code for *menopause present* (289903006)] **(C)**. Pirate plots of all females comparing survey and menopause general classifications **(D)**. Grey dashed lines in panel B and C indicate approximate menopause transition age range (40 to 60 years). Sample sizes ≤ 20 are obscured and reported as "20" in accordance with AoURP Data Dissemination Policy. All other participant counts were rounded up to nearest 5.

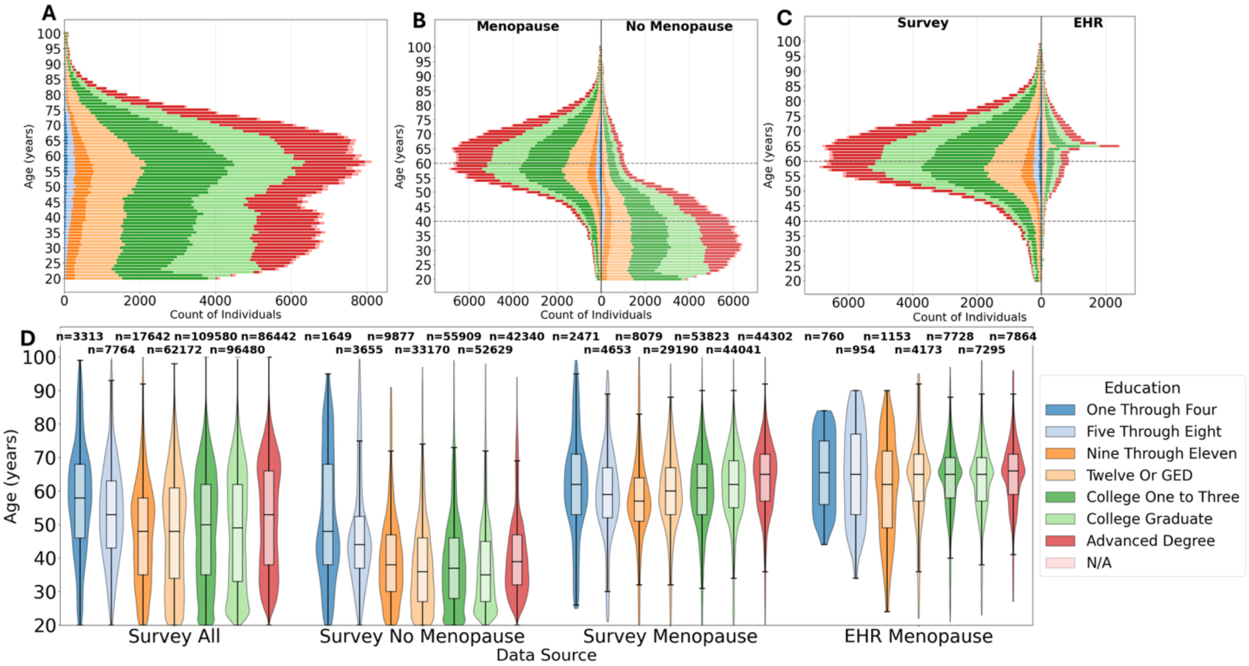


**Figure S8. AoURP v8 age distributions by education level.** Histograms with 1-year rounded bins for all female participants **(A)**, menopause survey response (answered "*yes none*" vs. "no" to "*Have your menstrual periods stopped permanently?*") **(B)**, and menopause survey response (answered "*yes none*" to "*Have your menstrual periods stopped permanently?*") vs. EHR menopause [SNOMED diagnostic code for *menopause present* (289903006)] **(C)**. Pirate plots of all females comparing survey and menopause general classifications **(D)**. Grey dashed lines in panels B and C indicate approximate menopause transition age range (40 to 60 years). Sample sizes ≤ 20 are obscured and reported as "20" in accordance with AoURP Data Dissemination Policy. All other participant counts were rounded up to nearest 5.

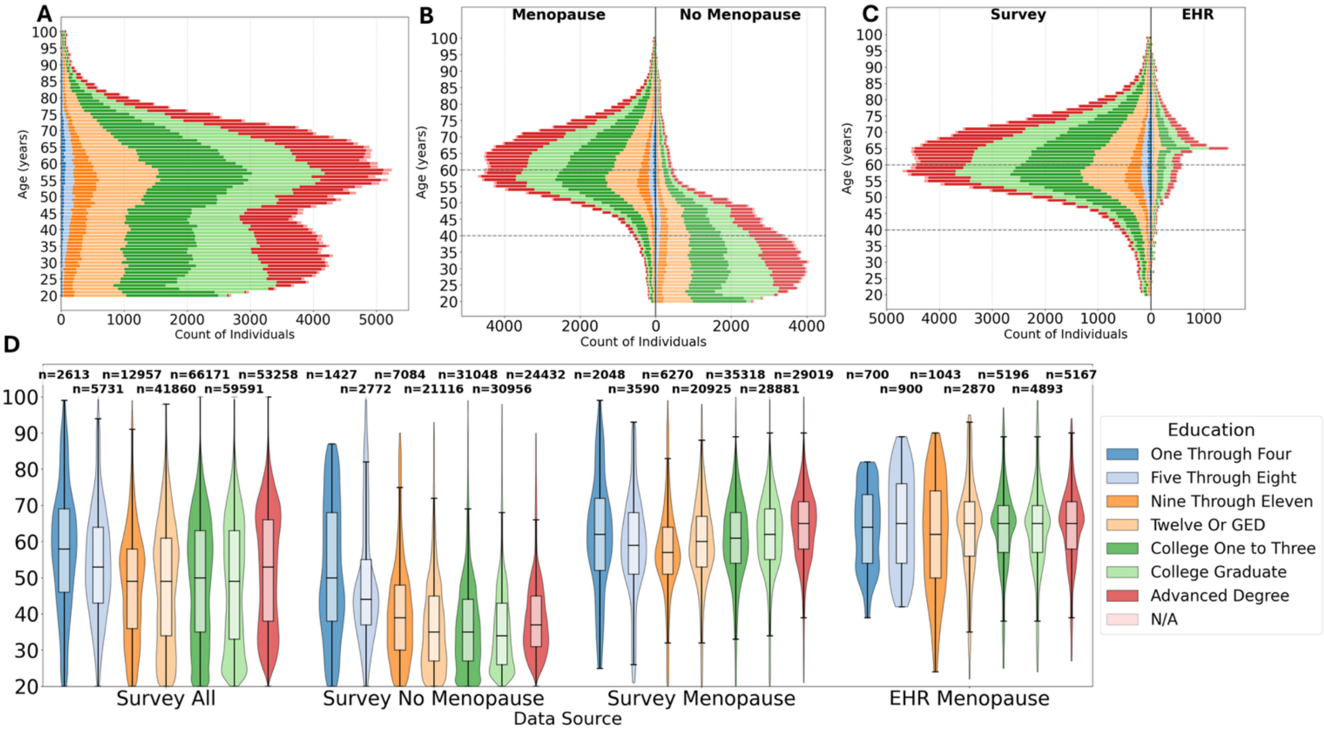


**Figure S9. AoURP v7 age distributions by education level.** Histograms with 1-year rounded bins for all female participants **(A)**, menopause survey response (answered "*yes none*" vs. "no" to "*Have your menstrual periods stopped permanently?*") **(B)**, and menopause survey response (answered "*yes none*" to "*Have your menstrual periods stopped permanently?*") vs. EHR menopause [SNOMED diagnostic code for *menopause present* (289903006)] **(C)**. Pirate plots of all females comparing survey and menopause general classifications **(D)**. Grey dashed lines in panels B and C indicate approximate menopause transition age range (40 to 60 years). Sample sizes ≤ 20 are obscured and reported as "20" in accordance with AoURP Data Dissemination Policy. All other participant counts were rounded up to nearest 5.

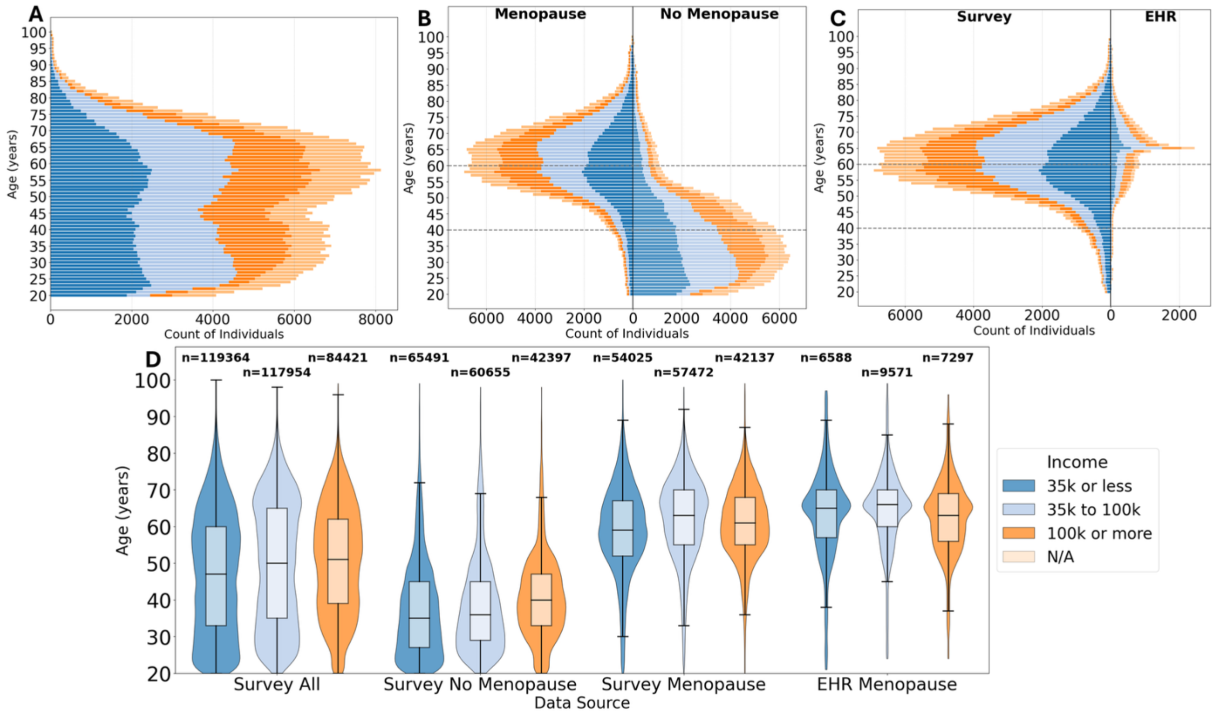


**Figure S10. AoURP v8 age distributions by income level (household) [combined into groups: $35,000 or less (35k or less), 35k to 100k, 100k or more, or not available (N/A)].** Histograms with 1-year rounded bins for all female participants **(A)**, menopause survey response (answered "*yes none*" vs. "no" to "*Have your menstrual periods stopped permanently?*") **(B)**, and menopause survey response (answered "*yes none*" to "*Have your menstrual periods stopped permanently?*") vs. EHR menopause [SNOMED diagnostic code for *menopause present* (289903006)] **(C)**. Pirate plots of all females comparing survey and menopause general classifications **(D)**. Grey dashed lines in panels B and C indicate approximate menopause transition age range (40 to 60 years). Sample sizes ≤ 20 are obscured and reported as "20" in accordance with AoURP Data Dissemination Policy. All other participant counts were rounded up to nearest 5.

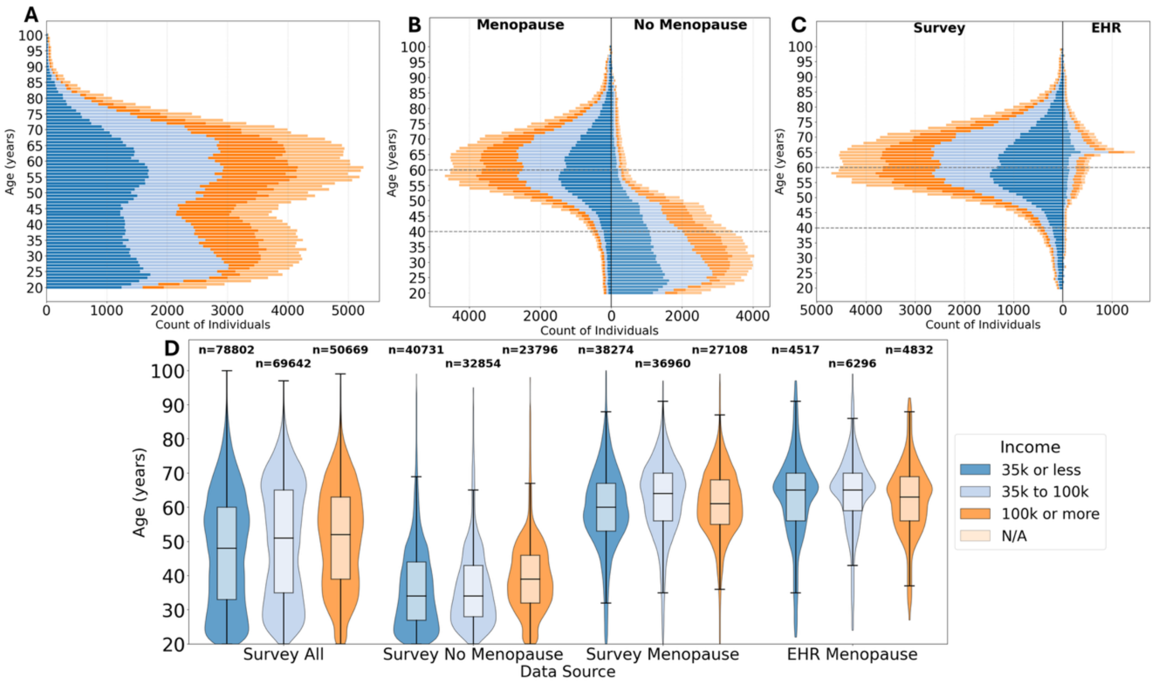


**Figure S11. AoURP v7 age distributions by income level.** Histograms with 1-year rounded bins for all female participants **(A)**, menopause survey response (answered "*yes none*" vs. "no" to "*Have your menstrual periods stopped permanently?*") **(B)**, and menopause survey response (answered "*yes none*" to "*Have your menstrual periods stopped permanently?*") vs. EHR menopause [SNOMED diagnostic code for *menopause present* (289903006)] **(C)**. Pirate plots of all females comparing survey and menopause general classifications **(D)**. Grey dashed lines in panels B and C indicate approximate menopause transition age range (40 to 60 years). Sample sizes ≤ 20 are obscured and reported as "20" in accordance with AoURP Data Dissemination Policy. All other participant counts were rounded up to nearest 5.

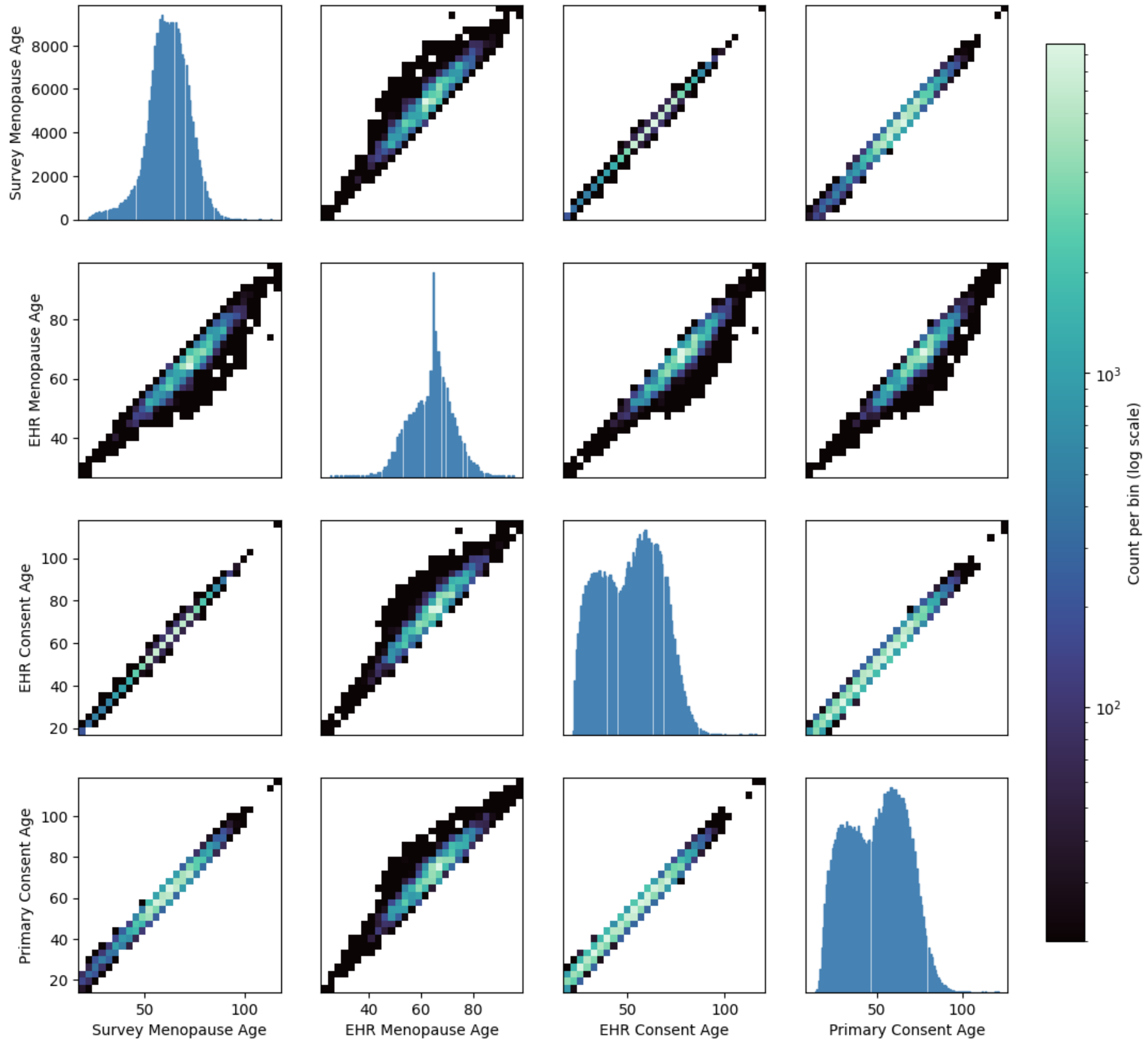


**Figure S12. Consistency between ages calculated from the survey and EHR in AoURP v7 for female participants.** Diagonal from left to right, top to bottom shows histograms for age at survey, age at EHR menopause note (note from earliest date/time for duplicate observations), EHR consent, and AoURP primary consent. Off diagonal plots show pairwise correlation heatmaps colored (black to light green color grid) by log (base 10) participant count per bin. Sample sizes ≤ 20 are obscured and reported as “20” in accordance with AoURP Data Dissemination Policy.
